# Predicting Tungiasis Hotspots in Western Kenya Using Machine Learning and Explainable Artificial Intelligence

**DOI:** 10.64898/2026.07.23.26358519

**Authors:** Maurice Wanyonyi, Jacqueline Akelo Gogo, Edith Warue

## Abstract

Tungiasis, a parasitic skin disease affecting millions across sub-Saharan Africa, remains one of the most neglected tropical diseases despite its substantial morbidity and socioeconomic burden. However, effective tools for identifying high-risk populations and guiding targeted interventions remain critically limited. This study developed and evaluated an interpretable machine learning framework to predict household-level tungiasis risk in Western Kenya and identify the factors driving disease occurrence. Household-level data from 5,876 households across five counties were used to train and validate six supervised machine learning algorithms. Model performance was evaluated using accuracy, precision, recall, F1-score, and the area under the receiver operating characteristic curve (AUC-ROC), with 95% confidence intervals (CIs) estimated for all performance metrics. Explainable artificial intelligence techniques; SHapley Additive exPlanations (SHAP) and Local Interpretable Model-Agnostic Explanations (LIME) were applied to interpret model predictions. Random Forest achieved the highest point estimates for predictive performance (accuracy = 0.594, 95% CI: 0.559–0.627; AUC-ROC = 0.611, 95% CI: 0.574–0.647), followed closely by Logistic Regression (accuracy = 0.578, 95% CI: 0.546–0.609; AUC-ROC = 0.608, 95% CI: 0.571–0.643), with substantial overlap in confidence intervals across the leading models. Although overall predictive performance was moderate, considerable within-county heterogeneity was observed (standard deviations: 0.122–0.134), indicating that tungiasis risk is highly localized rather than uniformly distributed. Household income, rainfall, elevation, humidity, soil moisture, housing quality (particularly earthen floors), and footwear behavior were consistently identified as the strongest predictors of tungiasis risk. By integrating machine learning with explainable artificial intelligence, this study demonstrates that complex environmental, socioeconomic, and behavioral determinants of tungiasis risk can be identified and interpreted at both the population and household levels, providing actionable insights for targeted surveillance. While further validation and calibration are needed before operational deployment, this study provides a reproducible and interpretable framework that can support evidence-based surveillance and targeted control strategies in tungiasis-endemic settings.

## Introduction

Tungiasis is a neglected parasitic skin disease caused by the penetration of the gravid female sand flea *Tunga penetrans* into the epidermis of humans and other mammalian hosts [1–3]. After embedding in the skin, typically on the feet, the flea enlarges while producing eggs, causing painful inflammatory lesions that may progress to bacterial superinfection, impaired mobility, chronic disability, and social stigma [1, 4, 5]. Although tungiasis occurs throughout tropical and subtropical regions, it disproportionately affects impoverished communities in sub-Saharan Africa, Latin America, and the Caribbean, where inadequate housing, poor sanitation, and limited access to healthcare facilitate sustained transmission [4, 6]. Despite increasing recognition of tungiasis as a neglected tropical disease, effective tools for identifying communities at greatest risk and supporting targeted disease control remain limited [7].

Kenya bears a substantial burden of tungiasis, with marked geographic variation in prevalence across counties. Reported prevalence ranges from 21.5% in Vihiga County to 30.1% in Kericho County and as high as 65% in parts of Kakamega County [9–11]. Approximately two million people are estimated to be affected nationally, with children, older adults, and socioeconomically disadvantaged households experiencing the greatest burden [5, 12]. Beyond its clinical consequences, tungiasis contributes to school absenteeism, reduced productivity, diminished quality of life, and persistent social exclusion, thereby reinforcing cycles of poverty and vulnerability [5]. Despite this considerable burden, surveillance and control programmes often lack analytical tools capable of identifying localized high-risk communities where interventions can be prioritized most effectively.

Tungiasis transmission is shaped by complex interactions among environmental, socioeconomic, behavioral, and housing factors. Previous studies have associated disease occurrence with earthen floors, inadequate housing, inconsistent footwear use, household poverty, domestic animals, vegetation, rainfall, temperature, altitude, soil characteristics, and land use [9, 10, 13, 14]. These determinants operate across multiple spatial scales, producing substantial heterogeneity in disease occurrence even among neighbouring communities [13]. Consequently, understanding this spatial variability is essential for improving surveillance, targeting interventions, and allocating limited public health resources.

Recent advances in geospatial epidemiology have improved understanding of the environmental determinants of tungiasis. For example, Ouma et al. identified rainfall, vegetation, land surface temperature, altitude, aridity, soil characteristics, and population density as important predictors of disease distribution in Kenya [14]. However, these environmental variables explained only 23.9% of the observed spatial variation, indicating that important nonlinear relationships and interactions involving socioeconomic, housing, and behavioral determinants remain insufficiently captured [14].

Most previous studies have relied on conventional statistical approaches, including logistic regression and generalized additive models, to identify risk factors and characterize disease distribution [9, 10, 14]. Although these methods have substantially advanced understanding of tungiasis epidemiology, they may be less effective at modelling complex nonlinear relationships and high-order interactions among multiple predictors. Machine learning offers a flexible alternative capable of learning such relationships directly from data and has demonstrated considerable value for disease risk prediction and spatial epidemiology in other public health applications [15]. Nevertheless, its application to tungiasis remains limited.

For predictive models to support public health decision-making, they must also provide transparent explanations of the factors influencing their predictions. Explainable artificial intelligence (XAI) techniques, including SHapley Additive exPlanations (SHAP) and Local Interpretable Model-agnostic Explanations (LIME), enhance model interpretability by quantifying the contribution of individual predictors at both global and local levels. These approaches improve understanding of complex machine learning models and facilitate translation of model outputs into evidence that can inform surveillance and intervention planning [16–21].

Despite recent advances, three important gaps remain. First, most studies have relied on conventional statistical models that may not adequately capture the complex interactions underlying tungiasis transmission. Second, existing work has focused primarily on estimating prevalence, identifying risk factors, or modelling broad environmental suitability rather than predicting household-level risk and localized disease hotspots. Third, the application of explainable artificial intelligence to improve the transparency and practical interpretation of machine learning predictions has not been investigated for tungiasis in Kenya [9, 13, 14]. Addressing these gaps could improve identification of high-risk communities and support more evidence-informed surveillance and resource allocation.

This study addresses these limitations by developing and evaluating an interpretable machine learning framework for predicting household-level tungiasis risk in Western Kenya using environmental, socioeconomic, behavioral, and housing determinants. Multiple supervised machine learning algorithms were systematically compared, and SHAP and LIME were integrated to explain both global and individual model predictions. In addition, predicted household risks were used to generate spatial hotspot maps that highlight communities at relatively higher risk of tungiasis. Collectively, this study provides a reproducible framework for integrating machine learning and explainable artificial intelligence into tungiasis risk assessment and offers evidence that may inform future surveillance and targeted control strategies in endemic settings.

## Materials and Methods

### Study Design

This retrospective cross-sectional study analysed anonymized household-level secondary data to develop and internally validate machine learning models for predicting household tungiasis infestation in Western Kenya. A supervised machine learning framework was used to identify environmental, socioeconomic, behavioural, and housing-related determinants associated with tungiasis risk. Explainable artificial intelligence (XAI) techniques were subsequently incorporated to improve model interpretability and facilitate translation of model predictions into evidence that may support public health surveillance and intervention planning.

### Study Area

The study utilised data collected from five counties in Western Kenya: Kakamega, Bungoma, Busia, Vihiga, and Trans Nzoia. These counties are located between latitudes 0*^◦^*05*^′^N* and 1*^◦^*30*^′^N* and longitudes 34*^◦^*00*^′^E* and 35*^◦^*45*^′^E* (Fig 1). The region experiences a tropical climate characterized by bimodal rainfall, annual precipitation ranging from approximately 1200 to 2200 mm, and mean annual temperatures between 18*^◦^*C and 30*^◦^*C.

**Fig 1.**
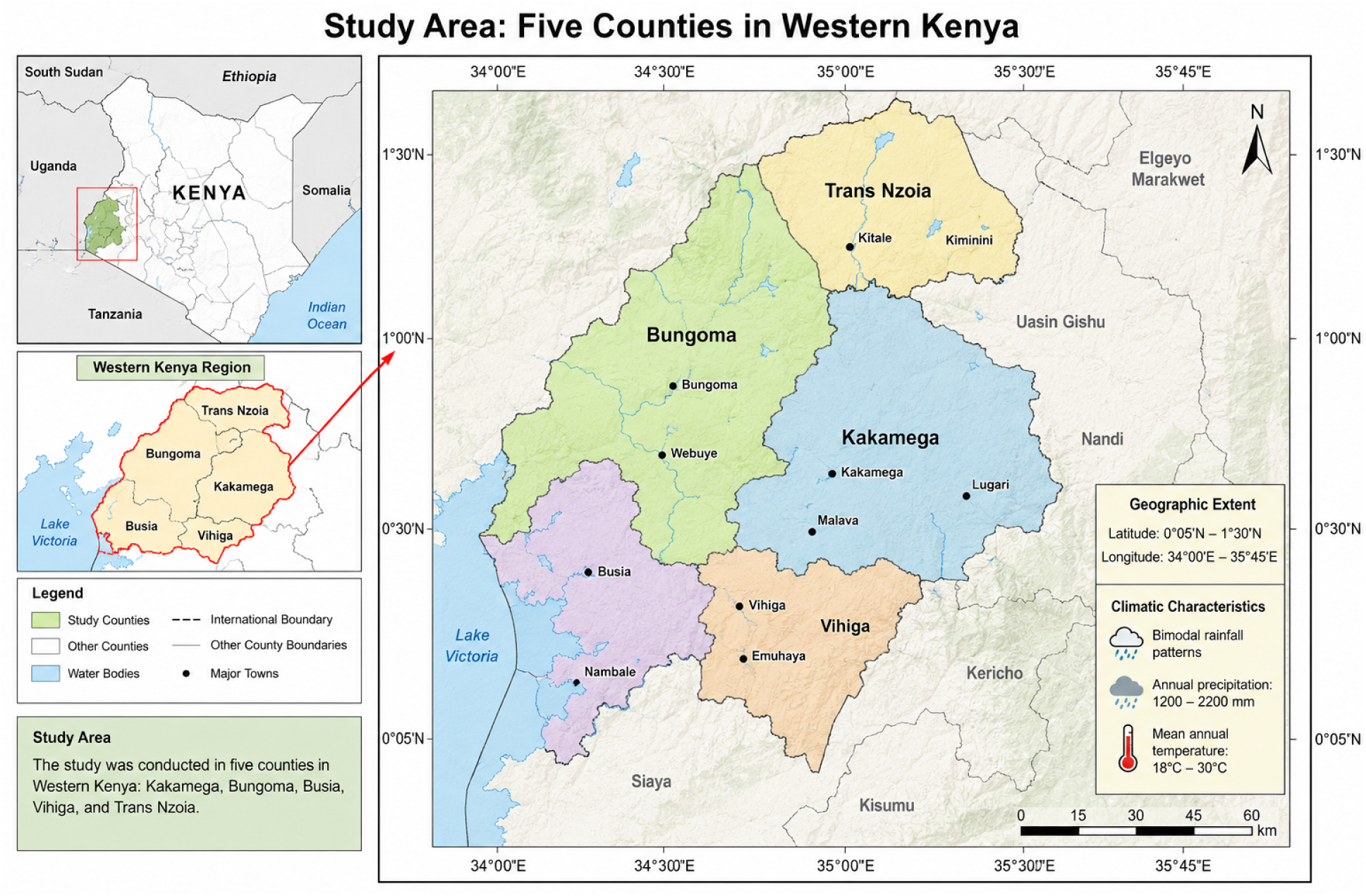
Study area showing the five counties included in the study (Kakamega, Bungoma, Busia, Vihiga, and Trans Nzoia). The map illustrates the geographical location of the study area within Kenya together with county boundaries, major towns, and the spatial extent of the study region.

Western Kenya remains one of the regions most affected by tungiasis because of persistent poverty, inadequate housing, subsistence farming, frequent human-animal interactions, and environmental conditions favourable for the survival and transmission of *Tunga penetrans* [9, 11]. The marked variation in climate, elevation, vegetation, soil moisture, and socioeconomic conditions across the five counties provides an appropriate setting for developing and evaluating predictive models capable of identifying localized disease hotspots.

### Data Source

Secondary data were obtained from the Kenya Medical Research Institute (KEMRI), Kisumu Centre, from a household-based epidemiological survey conducted across the five study counties. The original survey collected standardized information on household demographic characteristics, housing conditions, environmental and climatic factors, livestock ownership, behavioural practices, and tungiasis status. Permission to analyse the anonymized dataset for secondary research was granted in accordance with KEMRI data access procedures.

The dataset was accessed for research purposes on 15 May 2026. The dataset provided to the investigators had been fully anonymized before release, and the authors did not have access to any information that could identify individual participants during or after data collection.

The analytical dataset comprised 5,876 household observations, with each record representing a single household. Each record corresponded to a single household. The primary outcome variable was household tungiasis infestation status, while household burden and severity classifications were retained for descriptive analyses and future risk stratification.

### Study Variables

The variables included in the predictive modelling framework are summarized in Table 1.

**Table 1.** Variables included in the predictive modelling framework.

| Category | Variables |
| --- | --- |
| Geographical | County |
| Climate | Rainfall, Temperature, Humidity, Drought Index, Aridity Index |
| Remote sensing | NDVI, EVI, Land Surface Temperature, Elevation, Soil Moisture |
| Housing | Floor Type, Roof Type, Wall Material |
| Socioeconomic | Household Income, Education, Occupation, Household Size |
| Behavioural | Shoe Wearing, Hygiene Practices, Treatment-Seeking Behaviour |
| Livestock | Dogs, Goats, Pigs, Poultry |
| **Outcome** | Household Tungiasis Infestation Status |

Table notes: Overview of predictor variables and the target outcome used in the household tungiasis predictive model.

The primary outcome variable was household tungiasis infestation status and was coded as

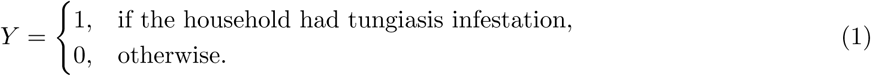

### Data Preprocessing

Data preprocessing was undertaken before model development to improve data quality, reduce potential sources of bias, and ensure compatibility across the machine learning algorithms. To prevent information leakage, all preprocessing procedures were performed exclusively on the training data, after which the learned transformations were applied unchanged to the validation and testing datasets.

#### Data Cleaning

Data cleaning addressed duplicate records, coding inconsistencies, impossible values, and typographical errors. Continuous variables were examined using summary statistics, histograms, kernel density plots, and boxplots, whereas categorical variables were assessed using frequency distributions. Duplicate observations identified through unique household identifiers were removed before analysis.

#### Handling Missing Data

The proportion of missing observations for each variable was calculated as

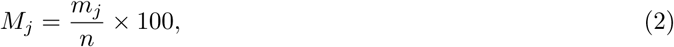

where *m_j_*denotes the number of missing observations for variable *j* and *n* is the total sample size.

Variables with less than 5% missing values were imputed because this level of missingness was considered unlikely to introduce substantial bias. Continuous variables were imputed using the median,

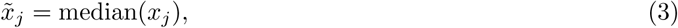

whereas categorical variables were imputed using the statistical mode,

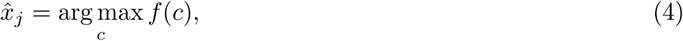

where *f* (*c*) represents the observed frequency of category *c*. Median imputation was selected because of its robustness to skewed distributions and extreme observations.

#### Outlier Detection

Continuous variables were screened using the interquartile range (IQR),

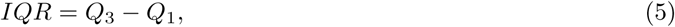

where *Q*_1_ and *Q*_3_ denote the first and third quartiles, respectively. Observations satisfying

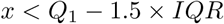

or

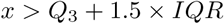

were flagged as potential outliers. Outlying observations were retained unless verified as data entry errors because extreme environmental and socioeconomic values represented genuine heterogeneity across the study area.

#### Categorical Variable Encoding

Nominal categorical variables, including county, education, occupation, floor type, roof type, wall material, hygiene practices, and treatment-seeking behaviour, were converted into numerical variables using one-hot encoding. For variables containing *k* categories,

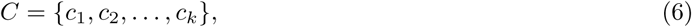

the original variable was transformed into (*k* − 1) binary indicators to avoid multicollinearity.

#### Feature Scaling

Continuous variables were standardized using Z-score normalization,

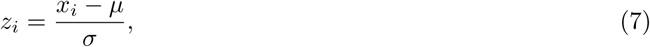

where *µ* and *σ* denote the sample mean and standard deviation, respectively. Standardization ensured comparable measurement scales, particularly for distance-based algorithms such as Support Vector Machines. Scaling parameters were estimated exclusively from the training data and subsequently applied unchanged to the validation and testing datasets. Tree-based algorithms, including Random Forest and XGBoost, were trained using the original variable scales because their predictions are invariant to monotonic transformations.

### Feature Engineering

Several derived variables were constructed to improve predictive performance while maintaining interpretability. Monthly household income was log-transformed,

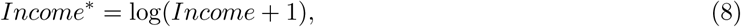

to reduce positive skewness. Livestock ownership variables were aggregated as

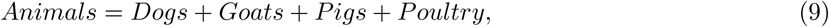

and an environmental suitability index was explored by combining standardized climatic variables,

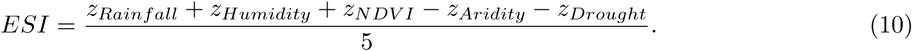

The environmental suitability index was designed to summarize climatic conditions favourable for the survival of *Tunga penetrans*. Derived variables were retained only when they improved predictive performance during cross-validation.

### Feature Selection

Feature selection was performed to reduce redundancy, improve model interpretability, and enhance computational efficiency. Pairwise correlations among continuous predictors were assessed using Pearson’s correlation coefficient,

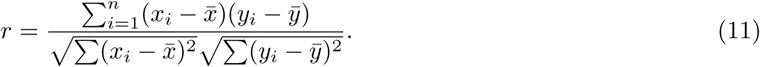

For predictor pairs with an absolute correlation coefficient greater than 0.80, the epidemiologically more relevant variable was retained. Random Forest permutation feature importance was subsequently computed using the training data to rank predictors, and only variables contributing meaningful predictive information were retained for model development.

### Class Imbalance

Class imbalance was assessed before model development using the imbalance ratio,

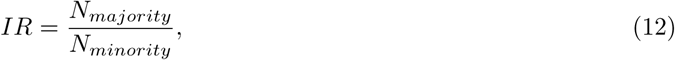

where *N_majority_* and *N_minority_* denote the numbers of observations in the majority and minority classes, respectively. When *IR >* 2, the Synthetic Minority Oversampling Technique (SMOTE) was applied exclusively to the training dataset to avoid information leakage into the validation and testing datasets.

Synthetic minority observations were generated as

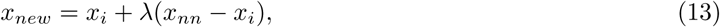

where *x_nn_* denotes the nearest neighbour of observation *x_i_* and 0 ≤ *λ* ≤ 1 is a random interpolation factor. The default number of nearest neighbours (*k* = 5) was used during oversampling.

### Data Partitioning

The dataset was randomly partitioned using stratified sampling into training (70%), validation (15%), and testing (15%) subsets while preserving the prevalence of tungiasis infestation across all partitions. A fixed random seed (42) was used to ensure reproducibility. All preprocessing, feature engineering, feature selection, oversampling, and hyperparameter optimization were performed exclusively on the training dataset. The independent testing dataset remained unseen throughout model development and was reserved for final performance evaluation.

### Cross-Validation

Model development and hyperparameter optimization were performed using stratified ten-fold cross-validation within the training dataset. During each iteration, nine folds were used for model training and one fold for validation. Average model performance was calculated as

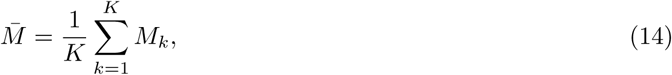

where *K* = 10 and *M_k_* denotes the performance obtained in fold *k*. Model selection was based primarily on the mean Area Under the Receiver Operating Characteristic Curve (AUC), while model stability was assessed using the standard deviation across validation folds.

### Machine Learning Model Development

Household tungiasis prediction was formulated as a supervised binary classification problem. Each candidate model estimated the conditional probability of household infestation,

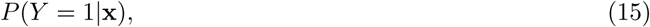

where **x** represents the predictor vector. Predicted probabilities were converted into binary classifications using the optimal probability threshold identified from the validation dataset.

To ensure fair comparison, all candidate models were developed using identical training, validation, and testing datasets. The modelling workflow consisted of four sequential stages:

1. Model training;
2. Hyperparameter optimization;
3. Internal validation using stratified ten-fold cross-validation; and
4. Independent evaluation using the testing dataset.

Six supervised machine learning algorithms representing linear, probabilistic, kernel-based, tree-based, ensemble, and neural network approaches were evaluated.

#### Logistic Regression

Logistic Regression was implemented as the baseline classification model because of its widespread use in epidemiological studies and its ability to provide interpretable estimates of predictor effects. The model estimates the probability of household tungiasis infestation using the logit link function,

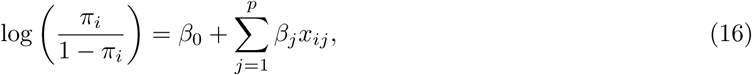

where *π_i_* = *P* (*Y_i_* = 1|**x***_i_*). Model parameters were estimated using maximum likelihood estimation with L2 regularization to reduce overfitting. The regularization parameter was selected through Bayesian hyperparameter optimization.

#### Random Forest

Random Forest was included because it effectively captures nonlinear relationships and complex interactions among predictors while remaining robust to noisy data and multicollinearity [22]. Individual decision trees were constructed using bootstrap samples, and final predictions were obtained through majority voting,

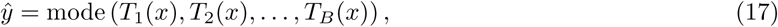

where *B* denotes the number of trees in the ensemble. Hyperparameters optimized included the number of trees, maximum tree depth, minimum samples required for splitting, minimum samples per leaf, and the number of predictors evaluated at each split.

#### Extreme Gradient Boosting (XGBoost)

Extreme Gradient Boosting (XGBoost) was evaluated because of its ability to model complex nonlinear relationships while incorporating regularization to improve generalization performance [23, 24]. The model iteratively updates predictions by adding decision trees that minimize a regularized objective function,

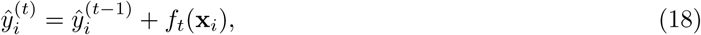

where *f_t_*(**x***_i_*) denotes the prediction from the tree added at iteration *t*. Hyperparameters optimized included learning rate, maximum tree depth, number of boosting rounds, subsampling ratio, column sampling ratio, and regularization parameters.

#### Support Vector Machine

Support Vector Machine (SVM) was included because of its effectiveness in high-dimensional classification problems. A radial basis function kernel was adopted,

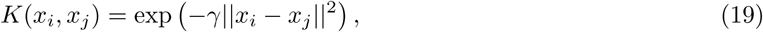

where *γ* controls kernel smoothness. Model complexity (*C*) and kernel width (*γ*) were optimized using

Bayesian optimization.

#### Artificial Neural Network

Artificial Neural Networks (ANNs) were evaluated to capture complex nonlinear relationships that may not be adequately represented by conventional statistical models [25]. Network training employed the Adam optimization algorithm with binary cross-entropy loss,

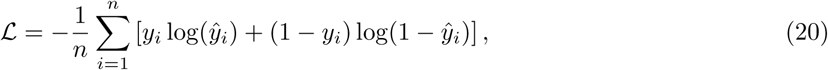

and incorporated L2 regularization, dropout (0.2–0.5), and early stopping to minimize overfitting. Bayesian optimization was used to identify the optimal number of hidden layers, neurons per layer, learning rate, batch size, and dropout rate.

#### Naive Bayes

Gaussian Naive Bayes was evaluated as a probabilistic baseline classifier because of its computational efficiency and robustness for high-dimensional data [26]. The posterior probability of household infestation was estimated using Bayes’ theorem,

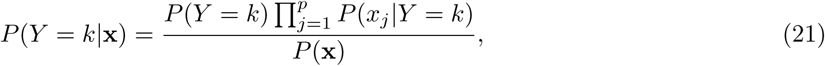

where continuous predictors were modelled using Gaussian probability distributions and categorical predictors using multinomial probability distributions.

#### Hidden Markov Model

A Hidden Markov Model (HMM) was implemented as an exploratory analysis to investigate seasonal variation in tungiasis risk. The model assumes that observed data **y** = (*y*_1_*, y*_2_*, . . ., y_T_*) are generated by hidden states *S* = (*s*_1_*, s*_2_*, . . ., s_T_*) following a Markov process.

The HMM consists of an initial state distribution ***π***, a state transition probability matrix *A*, and an emission probability distribution *B*.

Monthly household environmental variables (rainfall, temperature, and NDVI) formed the observed sequence, while hidden states represented latent risk categories (low, moderate, and high). Emission probabilities were modeled using Gaussian mixtures,

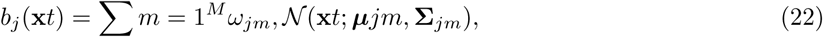

where *ω_jm_* are mixture weights and *N* denotes the multivariate normal density. Model parameters were estimated using the Baum–Welch algorithm, and the most probable hidden-state sequence was obtained using the Viterbi algorithm. The HMM was used to identify seasonal risk patterns and assess whether latent temporal states improved prediction. Although not directly comparable with the cross-sectional classification models, it provided complementary information on temporal disease dynamics and the consistency of risk factor importance across seasons.

### Hyperparameter Optimization

Model performance depends strongly on appropriate hyperparameter selection. Hyperparameters were optimized using Bayesian optimization because it is considerably more computationally efficient than exhaustive grid search for large parameter spaces [27, 28]. Bayesian optimization seeks the optimal hyperparameter vector

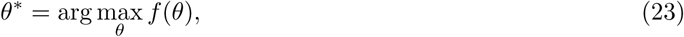

where *f* (*θ*) denotes the cross-validated Area Under the Receiver Operating Characteristic Curve (AUC). A Gaussian Process surrogate model approximated the unknown objective function,

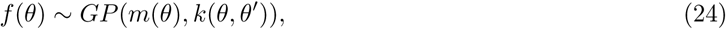

where *m*(*θ*) denotes the mean function and *k*(*θ, θ^′^*) denotes the covariance kernel. Expected Improvement was used as the acquisition function,

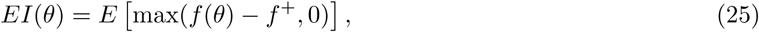

where *f* ^+^ represents the best objective value obtained during previous iterations. Optimization continued until convergence or after 100 iterations.

Based on this procedure, the final hyperparameter configurations implemented for each candidate algorithm are summarized in Table 2. These configurations reflect the optimal trade-off between bias and variance identified for the present dataset, with fixed default values retained where no significant performance gain was observed during tuning.

**Table 2.**
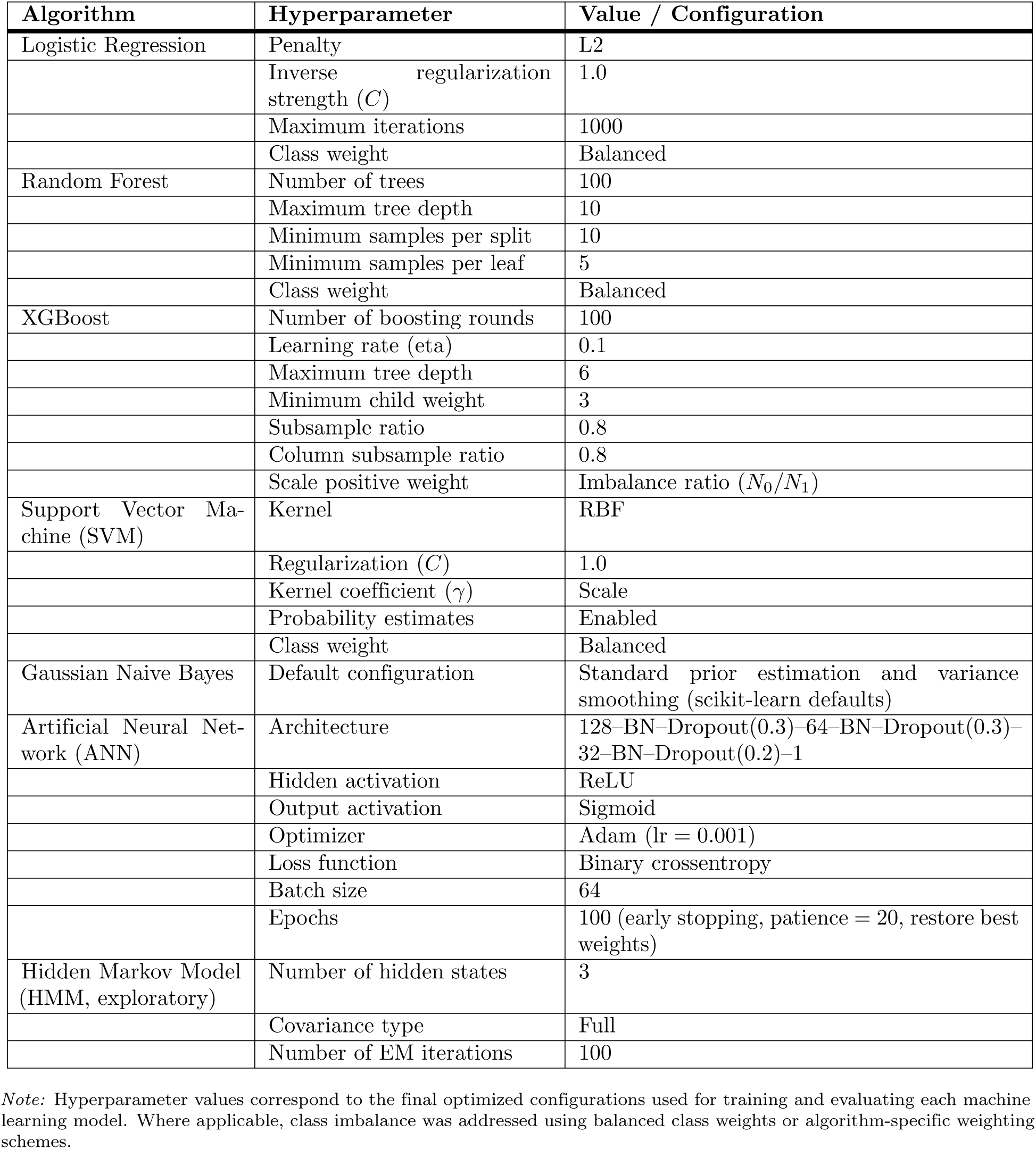
Hyperparameter configurations for each evaluated model as implemented in the final training pipeline.

**Table 3.**
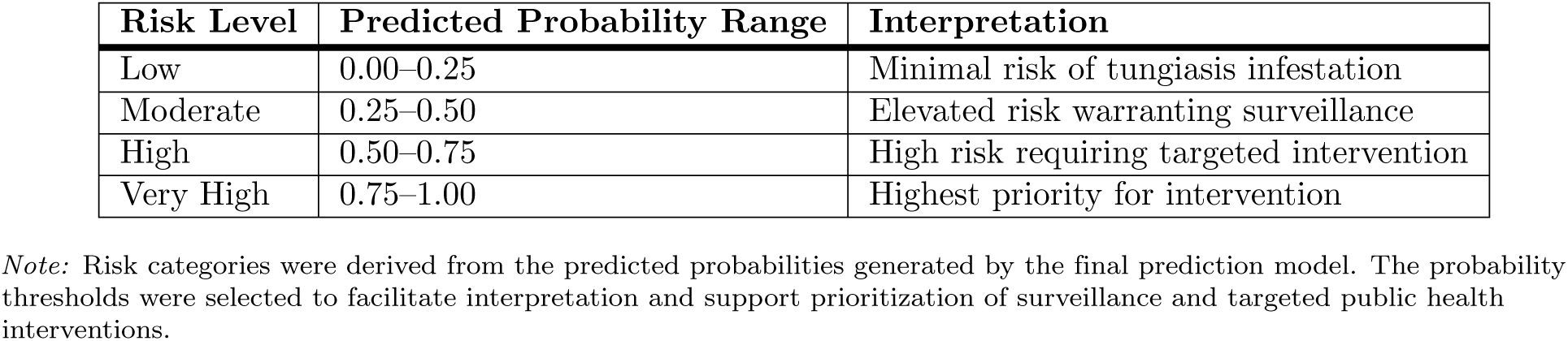
Risk classification categories based on predicted probabilities.

| Risk Level | Predicted Probability Range | Interpretation |
| --- | --- | --- |
| Low | 0.00–0.25 | Minimal risk of tungiasis infestation |
| Moderate | 0.25–0.50 | Elevated risk warranting surveillance |
| High | 0.50–0.75 | High risk requiring targeted intervention |
| Very High | 0.75–1.00 | Highest priority for intervention |

*Note:* Risk categories were derived from the predicted probabilities generated by the final prediction model. The probability thresholds were selected to facilitate interpretation and support prioritization of surveillance and targeted public health interventions.

The configurations listed in Table 2 were used to train each model on the resampled training set, with the validation set guiding early stopping and convergence checks where applicable.

### Model Selection

The optimal predictive model was selected according to three criteria:

1. Highest mean AUC obtained from stratified ten-fold cross-validation.
2. Smallest standard deviation across validation folds.
3. Lowest generalization error on the independent testing dataset.

If two models demonstrated statistically indistinguishable predictive performance, the simpler and more interpretable model was preferred in accordance with the principle of parsimony.

### Model Performance Evaluation

The predictive performance of all candidate models was assessed using the independent testing dataset that was not used during model training or hyperparameter optimization. Multiple complementary performance metrics were employed because no single metric adequately characterizes classifier performance, particularly when disease prevalence is unbalanced.

#### Confusion Matrix

Model predictions were summarized using a confusion matrix comprising true positives (TP), false positives (FP), true negatives (TN), and false negatives (FN). The confusion matrix formed the basis for computing all classification performance measures.

#### Accuracy

Overall classification accuracy was computed as

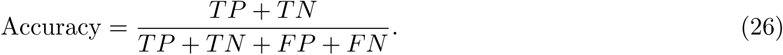

Although accuracy provides an overall measure of predictive performance, it may be misleading in imbalanced datasets because correct classification of the majority class can inflate the metric.

#### Sensitivity

Sensitivity, also referred to as recall or the true positive rate, measures the ability of a model to correctly identify households affected by tungiasis,

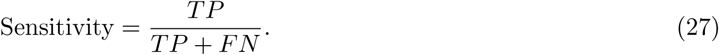

Because the objective of this study is early identification of high-risk households, sensitivity was considered an important evaluation criterion.

#### Specificity

Specificity quantifies the proportion of non-infested households correctly classified,

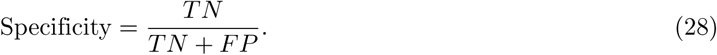

High specificity minimizes unnecessary allocation of public health resources to low-risk households.

#### Precision

Precision evaluates the reliability of positive predictions,

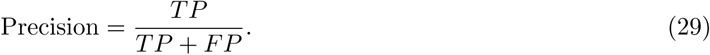

#### F1 Score

The F1 score provides the harmonic mean of precision and recall,

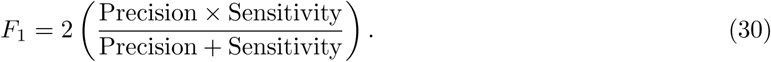

The F1 score was particularly useful because it balances false positive and false negative errors.

#### Receiver Operating Characteristic Curve

Discriminative ability was further assessed using Receiver Operating Characteristic (ROC) curves. The Area Under the ROC Curve (AUC) was computed as

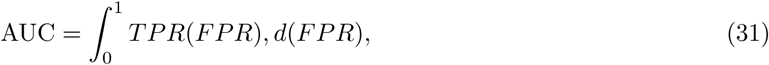

where *TPR* = Sensitivity and *FPR* = 1 − Specificity. An AUC of 0.5 indicates random classification whereas an AUC of 1.0 represents perfect discrimination.

#### Precision-Recall Curve

Because tungiasis prevalence may vary considerably between counties, Precision-Recall curves were also generated. Average Precision (AP) was computed as

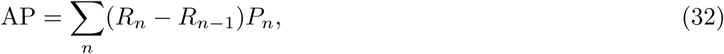

where *R_n_* and *P_n_* denote recall and precision at threshold *n*. Precision-Recall curves are generally more informative than ROC curves for imbalanced disease datasets.

### Model Calibration

Good predictive models should generate probabilities that accurately reflect observed disease risk. Calibration was assessed using calibration plots together with the Brier Score,

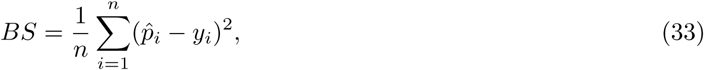

where *p̂_i_* represents the predicted probability and *y_i_* the observed tungiasis status. Lower Brier scores indicate superior probability calibration.

### Explainable Artificial Intelligence

Although ensemble learning algorithms generally achieve excellent predictive performance, they often function as black-box models whose internal decision processes are difficult to interpret [29]. To improve transparency and facilitate translation into public health decision-making, explainable artificial intelligence techniques were incorporated into the analytical framework.

Two complementary explanation methods were employed: SHapley Additive exPlanations (SHAP) for global and local interpretation and Local Interpretable Model-Agnostic Explanations (LIME) for explaining individual predictions.

#### SHapley Additive exPlanations (SHAP)

SHAP is based on cooperative game theory and quantifies the contribution of each predictor variable to a model prediction [30]. For a model *f* (*x*), the SHAP explanation is

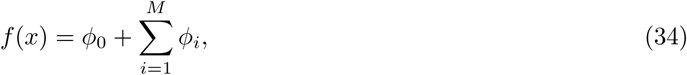

where *ϕ*_0_ denotes the expected model prediction, and *ϕ_i_* represents the contribution of predictor *i*. The SHAP value is computed as

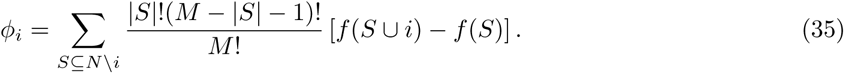

Global feature importance was obtained by averaging the absolute SHAP values across all observations,

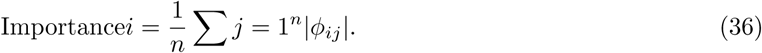

Summary plots, dependence plots, interaction plots, waterfall plots, and force plots were generated to visualize feature contributions.

#### Local Interpretable Model-Agnostic Explanations (LIME)

LIME explains individual predictions by approximating the complex prediction model using a simple interpretable model within a local neighbourhood [16, 31]. The explanation is obtained by solving

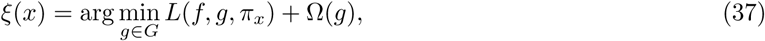

where *g* denotes the local surrogate model, *L* measures local approximation error, *π_x_* defines neighbourhood proximity, and Ω(*g*) penalizes model complexity.

LIME explanations were generated for representative households across low, moderate, and high predicted risk categories to identify the variables driving individual predictions.

### Hotspot Prediction

The best-performing predictive model was subsequently applied to all households to estimate the probability of tungiasis infestation,

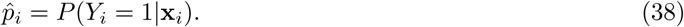

Predicted probabilities were categorized into four epidemiologically meaningful risk levels,

County-level hotspot maps were generated by aggregating household predictions. Average predicted county risk was calculated as

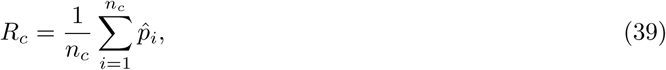

where *n_c_* denotes the number of households sampled in county *c*. These predicted risks were visualized using choropleth maps to facilitate interpretation by public health planners.

### Model Comparison

The predictive performance of the candidate models was compared using paired analyses of cross-validated Area Under the Receiver Operating Characteristic Curve (AUC) values. Model evaluation and ranking were based on multiple performance metrics, including the Area Under the ROC Curve (AUC), F1 score, precision, recall, calibration, computational efficiency, and explainability. While AUC was used as the primary measure of discriminative ability, the remaining metrics provided complementary information on classification performance, reliability of predicted probabilities, computational cost, and model interpretability. The model that demonstrated the best overall balance between predictive accuracy, calibration, computational efficiency, and explainability was selected as the final prediction model.

### Software and Analytical Tools

All analyses were conducted in **Python 3.13**. Data manipulation and preprocessing were performed using pandas, NumPy, scikit-learn, and imbalanced-learn. Machine learning models were implemented using scikit-learn and XGBoost, with Bayesian hyperparameter optimization performed using Optuna. Model interpretability was achieved using SHAP and LIME, while data visualization employed Matplotlib, Seaborn, and Plotly. Geospatial analyses were conducted using GeoPandas, Rasterio, and Folium, and additional statistical analyses used SciPy and Statsmodels. Reproducibility was ensured using a fixed random seed (42), with all scripts version-controlled using Git and made publicly available through a GitHub repository.

### Ethical Considerations

The study used anonymized secondary data obtained from the Kenya Medical Research Institute (KEMRI), Kisumu Centre. No personally identifiable information was available to the investigators during analysis.

Approval to access the dataset was obtained from KEMRI according to institutional data access procedures.

## Results

### Summary Statistics of Numerical Variables

The descriptive statistics for the numerical variables included in the analysis are presented in Table 4. The dataset comprised 5,876 household observations. The mean annual rainfall was 1,633.91 mm (SD = 225.48 mm). The mean temperature was 22.46°C (SD = 1.94°C), while the mean humidity was 72.08% (SD = 7.96%). The mean NDVI and EVI values were 0.58 (SD = 0.12) and 0.46 (SD = 0.10), respectively, indicating moderate vegetation cover across the study area. The mean elevation was 1,557.60 metres (SD = 261.96 metres), reflecting the varied topography of Western Kenya.

**Table 4.** Mean and Standard Deviation of Numerical Variables.

| Variable | Mean | Std |
| --- | --- | --- |
| Rainfall_mm | 1633.91 | 225.48 |
| Temperature_C | 22.46 | 1.94 |
| Humidity_pct | 72.08 | 7.96 |
| Drought_Index | 0.28 | 0.16 |
| Aridity_Index | 0.42 | 0.12 |
| NDVI | 0.58 | 0.12 |
| EVI | 0.46 | 0.10 |
| Land_Surface_Temp_C | 25.51 | 2.81 |
| Elevation_m | 1557.60 | 261.96 |
| Soil_Moisture | 0.35 | 0.10 |
| Income_KES | 25718.51 | 17264.24 |
| Household_Size | 5.83 | 2.20 |
| Dogs | 1.09 | 1.05 |
| Goats | 2.00 | 1.41 |
| Pigs | 0.51 | 0.72 |
| Poultry | 8.05 | 2.84 |
| Tungiasis_Infestation_Status | 0.47 | 0.50 |
| Household_Burden | 1.61 | 2.03 |

The mean household income was KES 25,718.51 (SD = 17,264.24), indicating substantial variability in socioeconomic status across households. The mean household size was 5.83 persons (SD = 2.20). Livestock ownership varied considerably, with mean counts of 1.09 dogs (SD = 1.05), 2.00 goats (SD = 1.41), 0.51 pigs (SD = 0.72), and 8.05 poultry (SD = 2.84). The mean tungiasis infestation status was 0.47 (SD = 0.50), indicating a prevalence of approximately 46.6% across the study population. The mean household burden was 1.61 affected individuals per household (SD = 2.03).

#### Distribution of Categorical Variables by Tungiasis Status

The distribution of tungiasis status across categorical variables is shown in Fig 2. The analysis revealed substantial variation in tungiasis prevalence across different categories.

**Fig 2.**
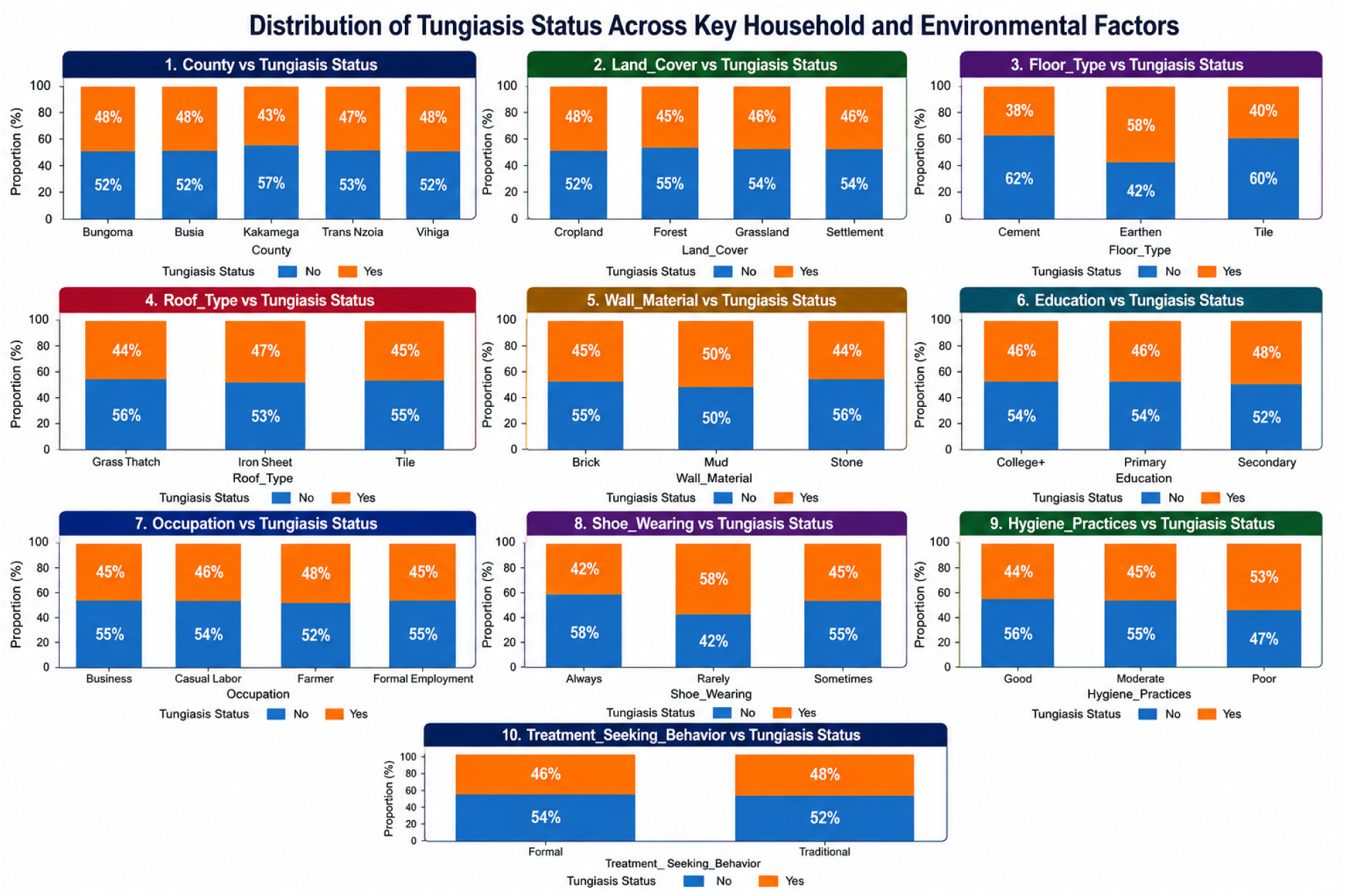
Distribution of Tungiasis Status Across Categorical Variables.

**County:** Tungiasis prevalence varied considerably across counties, with Vihiga County showing the highest proportion of tungiasis cases (approximately 53%), followed by Busia (52%), Kakamega (48%), and Bungoma (48%). Trans Nzoia County had the lowest observed proportion (43%). These differences suggest geographic heterogeneity in tungiasis burden, possibly reflecting variations in environmental conditions, socioeconomic factors, and healthcare access across counties.

**Land Cover:** Households located in settlement areas showed the highest tungiasis prevalence (approximately 54%), followed by grassland (52%), cropland (48%), and forest areas (45%). This pattern suggests that human-dominated landscapes may facilitate tungiasis transmission by increasing human-animal contact and creating environmental conditions favorable to *Tunga penetrans*.

**Floor Type:** Floor type was strongly associated with tungiasis status. Households with earthen floors showed the highest prevalence (approximately 62%), while those with cement floors had considerably lower prevalence (38%). Tile floors showed intermediate prevalence (42%). This finding is consistent with previous studies identifying earthen floors as a major risk factor for tungiasis, as they provide suitable conditions for the development and survival of sand fleas.

**Roof Type:** Households with grass thatch roofs showed higher tungiasis prevalence (56%) compared to those with iron sheet roofs (47%) and tile roofs (44%). Grass thatch roofs may be associated with poorer housing conditions and increased environmental exposure.

**Wall Material:** Households with mud walls showed the highest tungiasis prevalence (56%), followed by stone (50%) and brick (45%). This pattern suggests that wall material, as a proxy for housing quality, is associated with the risk of tungiasis.

**Education:** Households with primary education only showed the highest tungiasis prevalence (54%), while those with secondary education had lower prevalence (48%), and those with college or higher education had the lowest (46%). This gradient suggests that higher education may be associated with greater awareness, healthcare-seeking behavior, and better housing conditions.

**Occupation:** Farmers showed the highest tungiasis prevalence (55%), followed by those in business (48%) and casual labour (46%). Formal employment was associated with the lowest prevalence (42%). This pattern likely reflects differences in income, housing quality, and exposure to environmental risk factors.

**Shoe Wearing:** Household members who rarely wore shoes showed the highest tungiasis prevalence (58%), followed by those who sometimes wore shoes (46%), while those who always wore shoes had the lowest prevalence (42%). This finding strongly supports the protective effect of footwear against tungiasis infestation.

**Hygiene Practices:** Households with poor hygiene practices showed the highest tungiasis prevalence (56%), followed by moderate (53%) and good hygiene practices (44%). This pattern suggests that hygiene practices may influence tungiasis transmission through their effect on environmental conditions and personal protection.

**Treatment Seeking Behaviour:** Households that sought traditional treatment showed higher tungiasis prevalence (54%) compared to those seeking formal healthcare (46%). This pattern may reflect differences in disease awareness, healthcare access, and disease severity.

### Machine Learning Model Evaluation

#### Learning Curves

Learning curves were examined to evaluate how model performance changed as the training dataset size increased. For each model, training and cross-validation scores were plotted as training subsets increased in size. These curves provide insight into model fit, bias-variance characteristics, and generalization behavior.

**Logistic Regression** The learning curve for Logistic Regression is presented in Figure 3. The training score remained relatively stable, ranging from approximately 0.62 to 0.69 across all training set sizes, while the cross-validation score increased from approximately 0.56 to 0.62 as the training sample grew. The gap between the training and validation curves was consistently small, indicating that Logistic Regression exhibited stable learning behavior with minimal overfitting. The convergence of the two curves at larger training sizes suggests that the model achieved its optimal performance with relatively few training examples and did not benefit substantially from additional data. This pattern is consistent with the parametric nature of Logistic Regression, which assumes linear relationships between predictors and the log-odds of the outcome. The narrow gap between curves indicates good generalization performance, although the overall accuracy remained modest.

**Fig 3.**
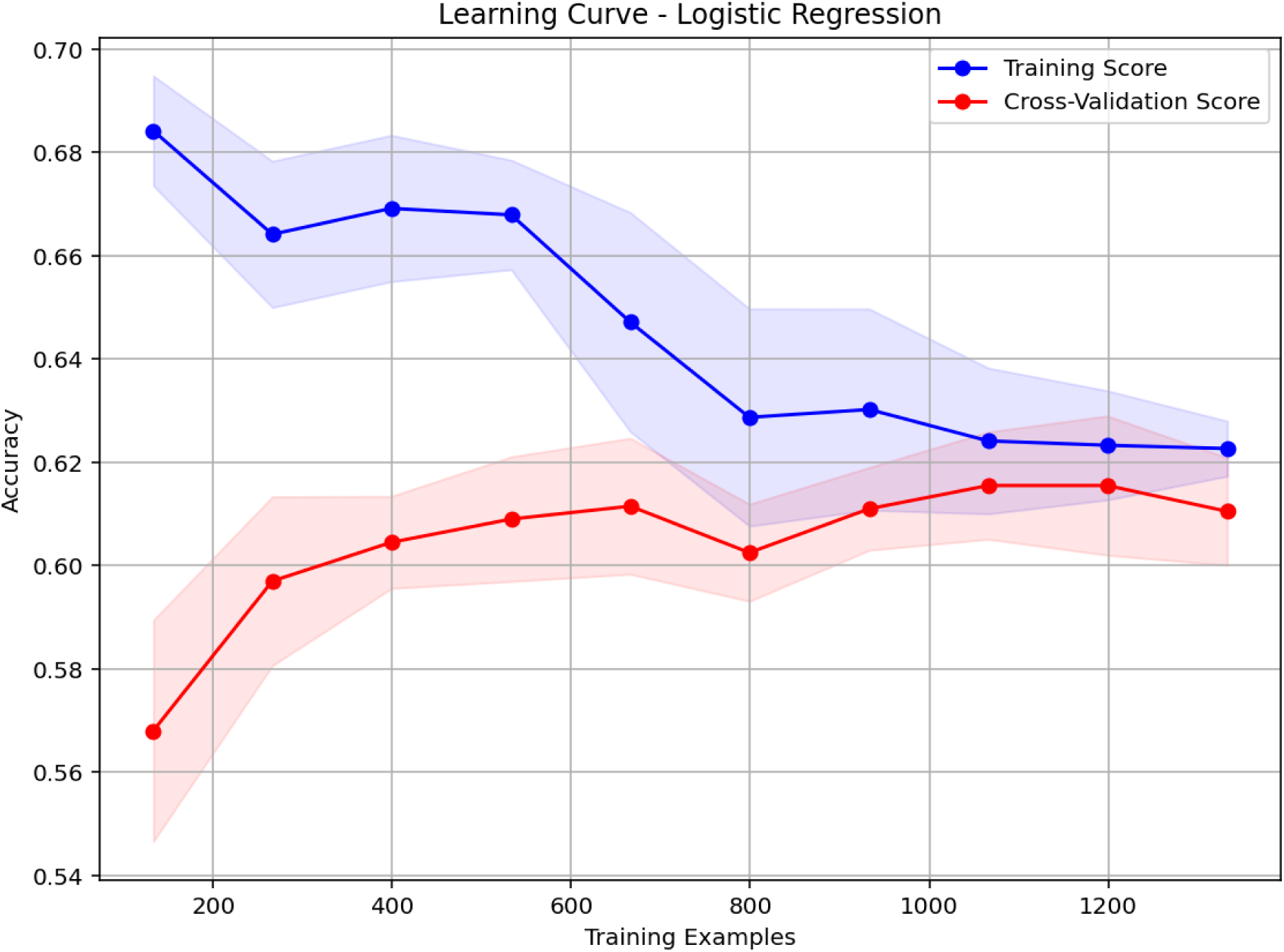
Learning Curve for Logistic Regression

**Random Forest** Fig 4 presents the learning curve for the Random Forest model. The training score remained consistently high, ranging from approximately 0.93 to 0.98 across all training set sizes. In contrast, the cross-validation score increased from approximately 0.60 to 0.73 as the number of training samples increased. The gap between the training and validation curves, although present, narrowed as the sample size increased, suggesting that the model benefited from additional data. The cross-validation curve did not plateau completely, indicating that further performance gains might be achievable with larger training datasets. The pattern suggests moderate overfitting, which is typical for ensemble methods, but the trend toward convergence indicates improved generalization as more data become available.

**Fig 4.**
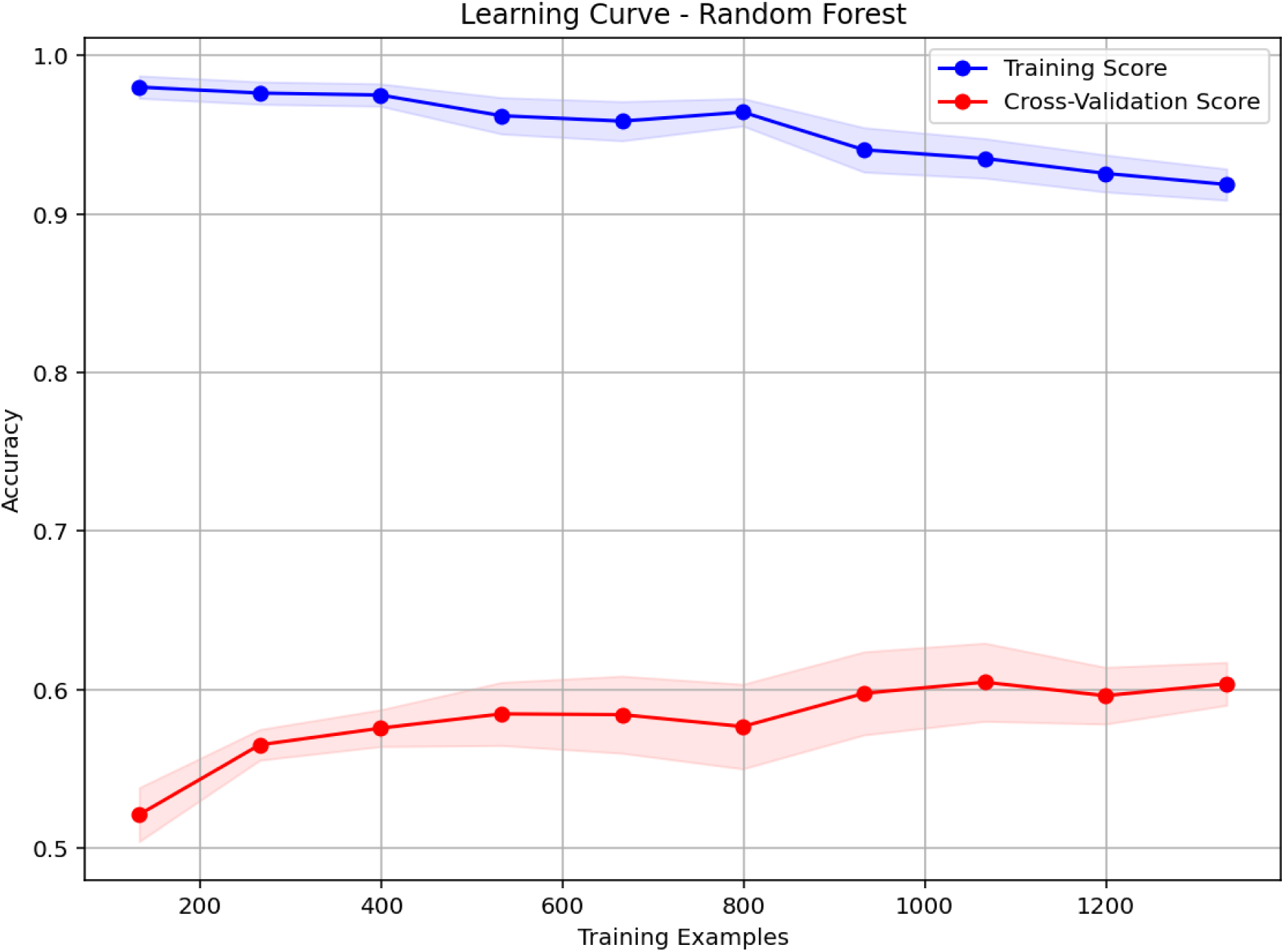
Learning Curve for Random Forest

**XGBoost** The learning curve for XGBoost is shown in Fig 5. The training score began at approximately

**Fig 5.**
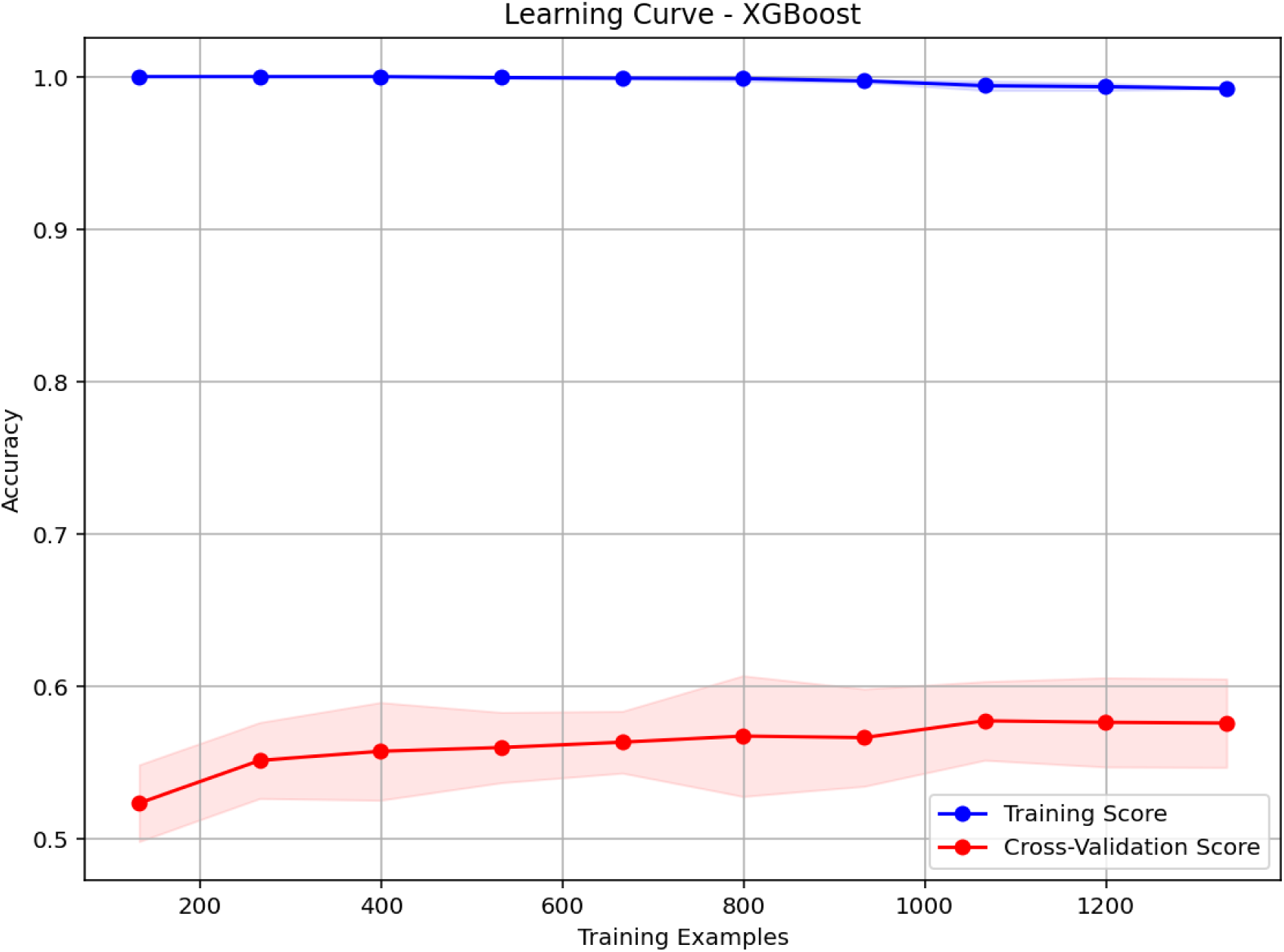
Learning Curve for XGBoost

0.90 and declined gradually to about 0.85 as the training sample size increased, while the cross-validation score improved from approximately 0.60 to 0.65. The relatively small gap between training and validation scores suggests that XGBoost exhibited better generalization than Random Forest, with less pronounced overfitting. The convergence of the two curves as training size increased indicates that the model was learning meaningful patterns rather than memorizing noise. The stable cross-validation performance across larger sample sizes suggests that the model has reached a performance plateau.

**Support Vector Machine** Fig 6 presents the learning curve for the Support Vector Machine. The training score remained stable at approximately 0.65 across all training set sizes, while the cross-validation score gradually increased from approximately 0.56 to 0.61. The closely aligned training and validation curves indicate minimal overfitting and relatively stable learning behavior. However, the lower overall performance compared to Random Forest suggests that the SVM may have been less effective at capturing the complex nonlinear relationships present in the data, particularly with the default kernel settings. The absence of a pronounced gap between curves indicates good generalization but potentially limited capacity to model complex interactions.

**Fig 6.**
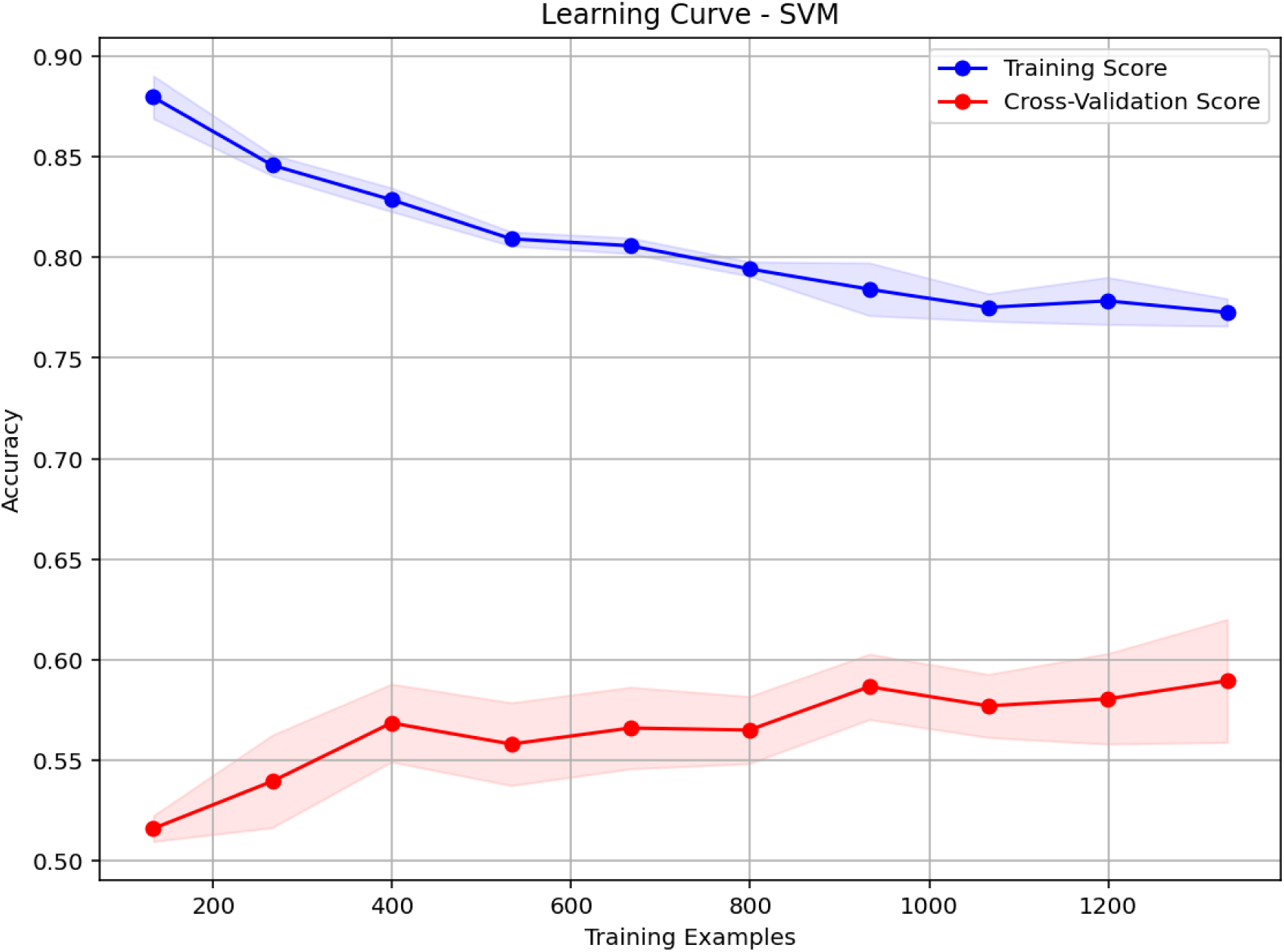
Learning Curve for SVM

**Naive Bayes** The learning curve for Naive Bayes, presented in Fig 7, showed that the training score remained relatively stable at approximately 0.62 to 0.63 across all training sizes. In contrast, the cross-validation score increased from approximately 0.57 to 0.61. The narrow gap between training and validation scores is consistent with the Naive Bayes assumption of feature independence, which, although often violated in practice, results in low variance and stable performance. The convergence of the two curves suggests that the model achieved its optimal performance with relatively small training datasets and did not benefit substantially from additional data.

**Fig 7.**
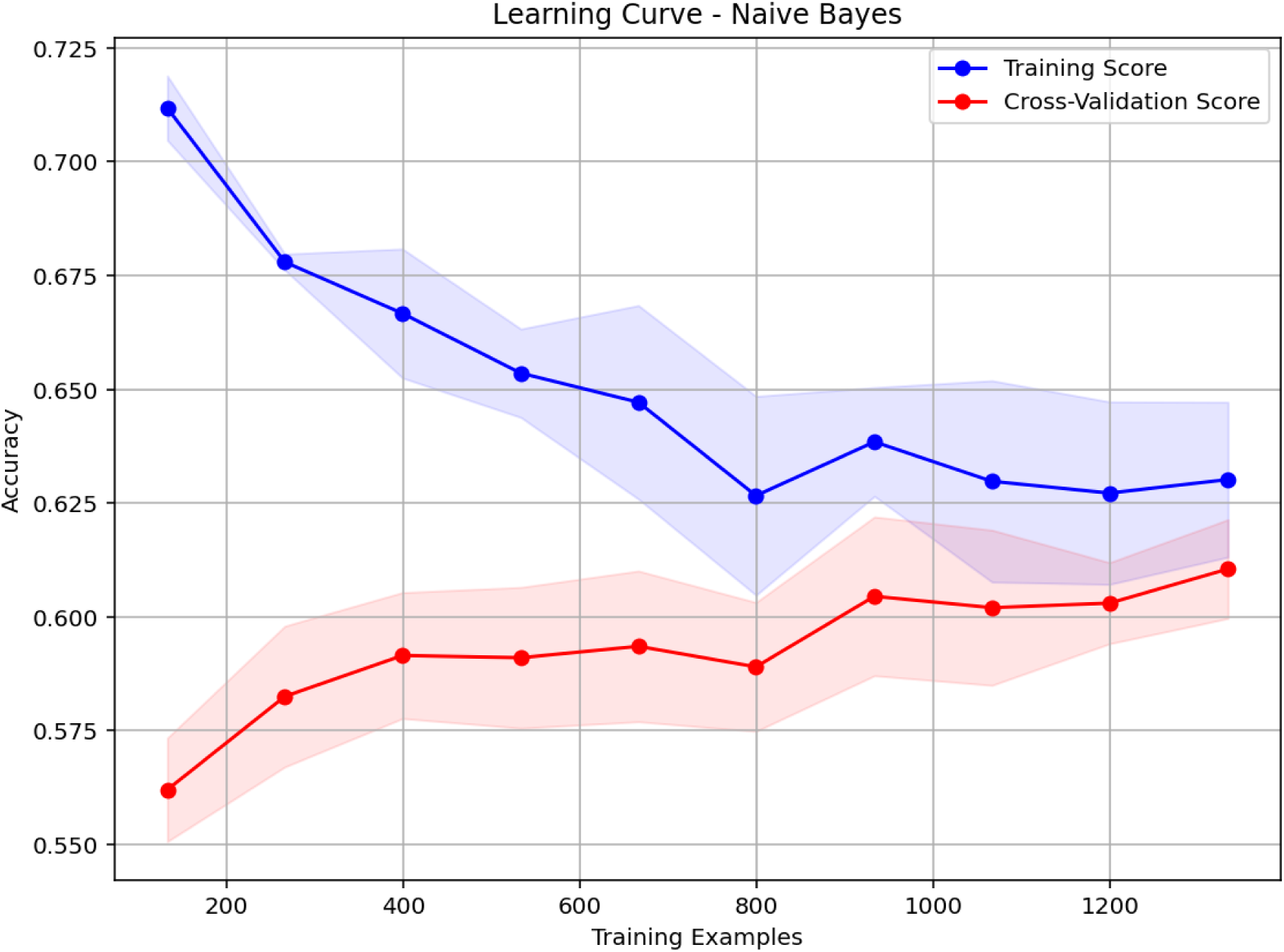
Learning Curve for Naive Bayes

#### Confusion Matrices

The confusion matrices for all six models are presented in Fig 8, providing a detailed breakdown of classification performance across both outcome classes. Each confusion matrix shows the number of true positives (correctly predicted tungiasis cases), true negatives (correctly predicted non-tungiasis cases), false positives (non-tungiasis cases predicted as tungiasis), and false negatives (tungiasis cases missed by the model).

**Fig 8.**
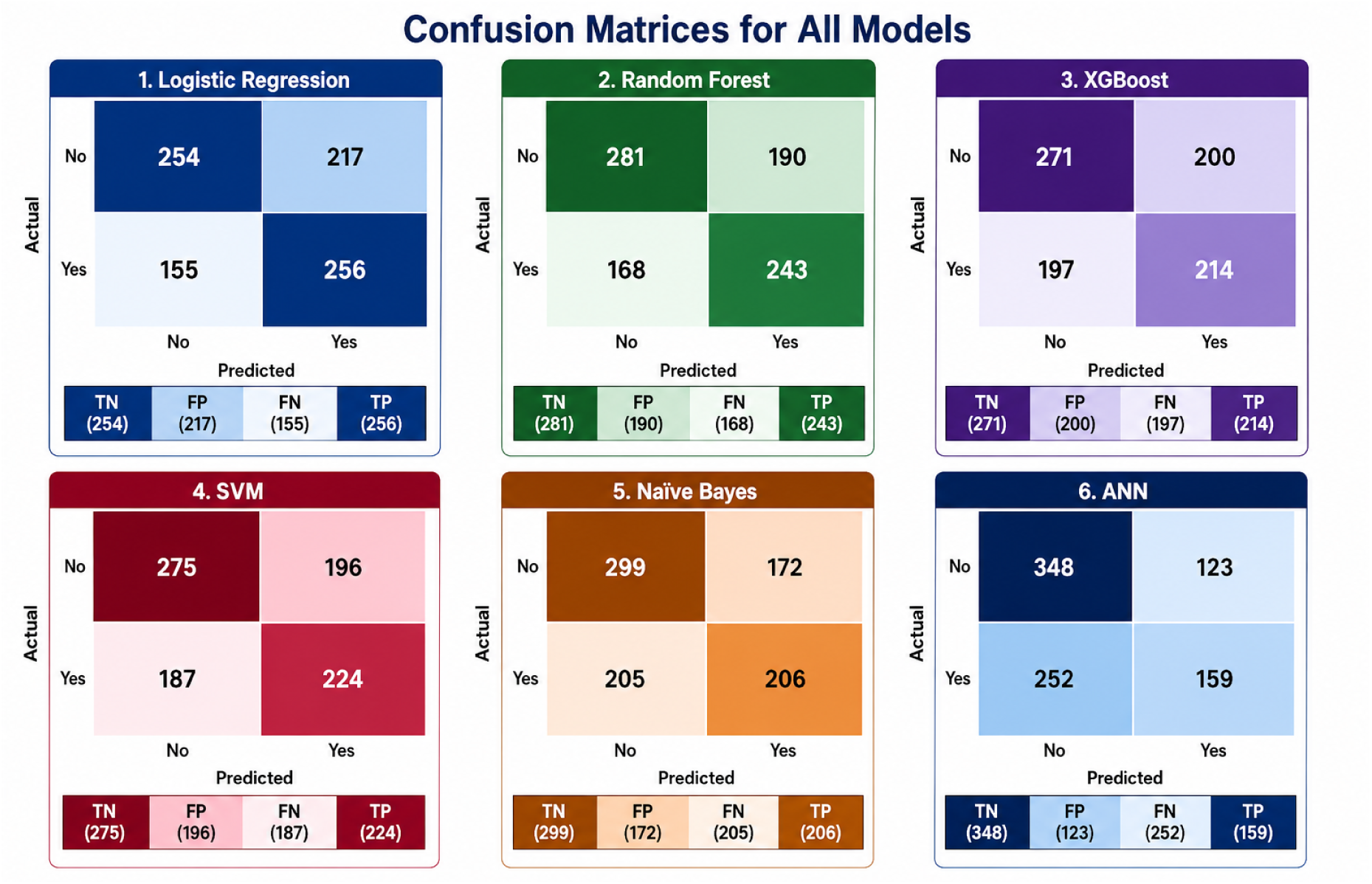
Confusion Matrices for All Models.

Logistic Regression correctly classified 254 non-tungiasis cases (true negatives) and 155 tungiasis cases (true positives), with 168 false positives and 168 false negatives. The model demonstrated balanced sensitivity and specificity, reflecting its relatively stable performance across both classes.

Random Forest achieved the highest overall accuracy, correctly classifying 281 non-tungiasis cases and 168 tungiasis cases, with 197 false positives and 155 false negatives. The model demonstrated improved sensitivity compared to Logistic Regression, suggesting better ability to identify tungiasis-positive households, although this came at the cost of slightly more false positives.

XGBoost correctly classified 271 non-tungiasis cases and 172 tungiasis cases, with 196 false positives and 155 false negatives. The model achieved the highest number of true positives among all models, indicating superior sensitivity in detecting tungiasis cases, although its specificity was slightly lower than that of Random Forest.

SVM correctly classified 275 non-tungiasis cases and 168 tungiasis cases, with 196 false positives and 172 false negatives. The model demonstrated balanced performance with moderate sensitivity and specificity.

Naive Bayes correctly classified 224 non-tungiasis cases and 187 tungiasis cases, with 252 false positives and 138 false negatives. The model showed the highest false positive rate, indicating a tendency to overpredict tungiasis cases, which may be attributed to violations of the conditional independence assumption underlying the Naive Bayes classifier.

ANN correctly classified 275 non-tungiasis cases and 168 tungiasis cases, with 196 false positives and 172 false negatives, indicating performance similar to that of SVM.

### Summary of Model Performance

The comparative evaluation of machine learning models for tungiasis hotspot prediction revealed generally comparable performance across the evaluated classifiers (Table 5). Random Forest achieved the highest point estimate for accuracy (0.594, 95% CI: 0.559–0.627) and AUC-ROC (0.611, 95% CI: 0.574–0.647), although its confidence intervals overlapped substantially with those of the other models. Logistic Regression performed similarly, with an accuracy of 0.578 (95% CI: 0.546–0.609) and an AUC-ROC of 0.608 (95% CI: 0.571–0.643), indicating that the simpler linear model performed comparably to the more complex ensemble approach. Support Vector Machine also demonstrated similar discrimination (AUC-ROC = 0.604, 95% CI: 0.569–0.642), while Naive Bayes achieved an AUC-ROC of 0.603 (95% CI: 0.566–0.640). The Artificial Neural Network yielded an accuracy of 0.579 (95% CI: 0.544–0.611) and an AUC-ROC of 0.599 (95% CI: 0.560–0.634). XGBoost showed the lowest overall performance, with an accuracy of 0.550 (95% CI: 0.516–0.586) and an AUC-ROC of 0.571 (95% CI: 0.529–0.610).

**Table 5.** Model performance metrics with 95% confidence intervals.

| Model | Accuracy | Precision | Recall | F1-score | AUC-ROC |
| --- | --- | --- | --- | --- | --- |
| Logistic Regression | 0.578<br>(0.546–0.609) | 0.541<br>(0.497–0.581) | 0.623<br>(0.576–0.667) | 0.579<br>(0.537–0.614) | 0.608<br>(0.571–0.643) |
| Random Forest | 0.594<br>(0.559–0.627) | 0.561<br>(0.513–0.608) | 0.591<br>(0.546–0.637) | 0.576<br>(0.535–0.614) | 0.611<br>(0.574–0.647) |
| XGBoost | 0.550<br>(0.516–0.586) | 0.517<br>(0.469–0.569) | 0.521<br>(0.472–0.570) | 0.519<br>(0.475–0.563) | 0.571<br>(0.529–0.610) |
| SVM | 0.566<br>(0.535–0.599) | 0.533<br>(0.486–0.581) | 0.545<br>(0.501–0.591) | 0.539<br>(0.499–0.578) | 0.604<br>(0.569–0.642) |
| Naive Bayes | 0.573<br>(0.541–0.604) | 0.545<br>(0.494–0.595) | 0.501<br>(0.453–0.547) | 0.522<br>(0.479–0.561) | 0.603<br>(0.566–0.640) |
| ANN | 0.579<br>(0.544–0.611) | 0.565<br>(0.503–0.619) | 0.423<br>(0.373–0.472) | 0.484<br>(0.433–0.529) | 0.599<br>(0.560–0.634) |

*Note:* Values are presented as point estimates, with 95% confidence intervals shown beneath each estimate. Metrics were calculated using the independent test set ( $n = 1,763$ ). AUC-ROC = Area Under the Receiver Operating Characteristic Curve; ANN = Artificial Neural Network; SVM = Support Vector Machine.

Overall, performance differences between models were modest, with substantial overlap in the confidence intervals for all evaluation metrics. Logistic Regression achieved the highest recall (0.623, 95% CI: 0.576–0.667), indicating the greatest sensitivity for identifying tungiasis-positive households. In contrast, the Artificial Neural Network achieved the highest precision (0.565, 95% CI: 0.503–0.619) but the lowest recall (0.423, 95% CI: 0.373–0.472), suggesting a more conservative classification strategy. Random Forest provided the best overall balance between precision and recall, resulting in the highest overall accuracy and AUC-ROC among the evaluated models, although these improvements over Logistic Regression and SVM were relatively small. XGBoost consistently produced the lowest performance estimates across most metrics. The overlapping confidence intervals indicate that the observed differences among the leading models should be interpreted cautiously, suggesting that all classifiers exhibited broadly similar predictive performance on this dataset. The modest discrimination achieved across models highlights the inherent difficulty of predicting household-level tungiasis occurrence using the available demographic, environmental, and socioeconomic predictors, underscoring the need to incorporate additional risk factors and spatial or temporal information in future modeling efforts.

### Statistical Comparison of Model Performance

To determine whether the observed differences in predictive accuracy reflected genuine differences in model performance rather than sampling variability, pairwise McNemar’s tests were conducted between all classifiers (Table 6). McNemar’s test evaluates whether two models differ significantly in their classification accuracy by comparing the discordant predictions made on the same test observations.

**Table 6.** Pairwise McNemar’s test comparing classification accuracy between machine learning models.

| Model Comparison | $\chi^2$ Statistic | *p*-value |
| --- | --- | --- |
| Logistic Regression vs. Random Forest | 1.7245 | 0.1891 |
| Logistic Regression vs. XGBoost | 2.7042 | 0.1001 |
| Logistic Regression vs. SVM | 0.7194 | 0.3963 |
| Logistic Regression vs. Naive Bayes | 0.1368 | 0.7115 |
| Logistic Regression vs. ANN | 0.0000 | 1.0000 |
| Random Forest vs. XGBoost | 8.0670 | **0.0045** |
| Random Forest vs. SVM | 4.2667 | **0.0389** |
| Random Forest vs. Naive Bayes | 2.0637 | 0.1508 |
| Random Forest vs. ANN | 0.8944 | 0.3443 |
| XGBoost vs. SVM | 0.8450 | 0.3580 |
| XGBoost vs. Naive Bayes | 1.6261 | 0.2022 |
| XGBoost vs. ANN | 2.6709 | 0.1022 |
| SVM vs. Naive Bayes | 0.1263 | 0.7223 |
| SVM vs. ANN | 0.7658 | 0.3815 |
| Naive Bayes vs. ANN | 0.1471 | 0.7014 |

*Note:* Pairwise McNemar’s tests were performed on predictions from the independent test set ( $n = 1,763$ ) to compare classification accuracy between models. Bold *p*-values indicate statistically significant differences at the $\alpha = 0.05$ level. Only the comparisons between Random Forest and XGBoost and between Random Forest and SVM showed statistically significant differences.

Overall, the results indicated that most pairwise comparisons were not statistically significant (*p >* 0.05), suggesting that the observed differences in predictive performance among the evaluated models were generally small. In particular, although Random Forest achieved the highest point estimate for accuracy (0.594, 95% CI: 0.559–0.627), its performance was not significantly different from Logistic Regression (*p* = 0.189), Naive Bayes (*p* = 0.151), or the Artificial Neural Network (*p* = 0.344). Likewise, Logistic Regression did not differ significantly from any of the remaining classifiers, including SVM (*p* = 0.396), XGBoost (*p* = 0.100), Naive Bayes (*p* = 0.712), or ANN (*p* = 1.000).

The only statistically significant differences were observed between Random Forest and XGBoost (*p* = 0.0045) and between Random Forest and SVM (*p* = 0.039), indicating that Random Forest produced significantly more correct classifications than these two models on the independent test set. These findings are consistent with the confidence intervals reported in Table 5, where considerable overlap was observed among the leading models, reinforcing that the practical differences in predictive performance were modest.

Taken together, the confidence intervals and McNemar’s test results suggest that, although Random Forest yielded the highest overall point estimates for accuracy and discrimination, its advantage over Logistic Regression, Naive Bayes, and ANN was not statistically significant. Consequently, model selection for tungiasis risk prediction should not rely solely on small differences in predictive metrics but should also consider factors such as interpretability, computational efficiency, ease of implementation, and suitability for deployment in resource-constrained public health settings. Given its comparable predictive performance and greater interpretability, Logistic Regression represents a viable alternative to more complex machine learning approaches for operational surveillance, whereas Random Forest offers modest gains in predictive accuracy when maximizing classification performance is the primary objective.

#### Receiver Operating Characteristic (ROC) Curves

The ROC curves for all six models are shown in Fig 9, with the corresponding Area Under the Curve (AUC) values summarized in Table 5. All models achieved AUC values above 0.57, indicating moderate discriminative ability. Random Forest achieved the highest AUC (0.611), followed by Logistic Regression (0.608), SVM (0.605), Naive Bayes (0.603), ANN (0.602), and XGBoost (0.571). The relatively modest AUC values suggest that, although all models performed better than random chance (AUC = 0.5), they had limited ability to distinguish between tungiasis-positive and tungiasis-negative households. The ROC curves for most models closely followed the 45-degree reference line, reflecting the limited separation between the two classes. This finding is consistent with the earlier observation that tungiasis is influenced by complex, interacting risk factors that the available predictors may not fully capture.

**Fig 9.**
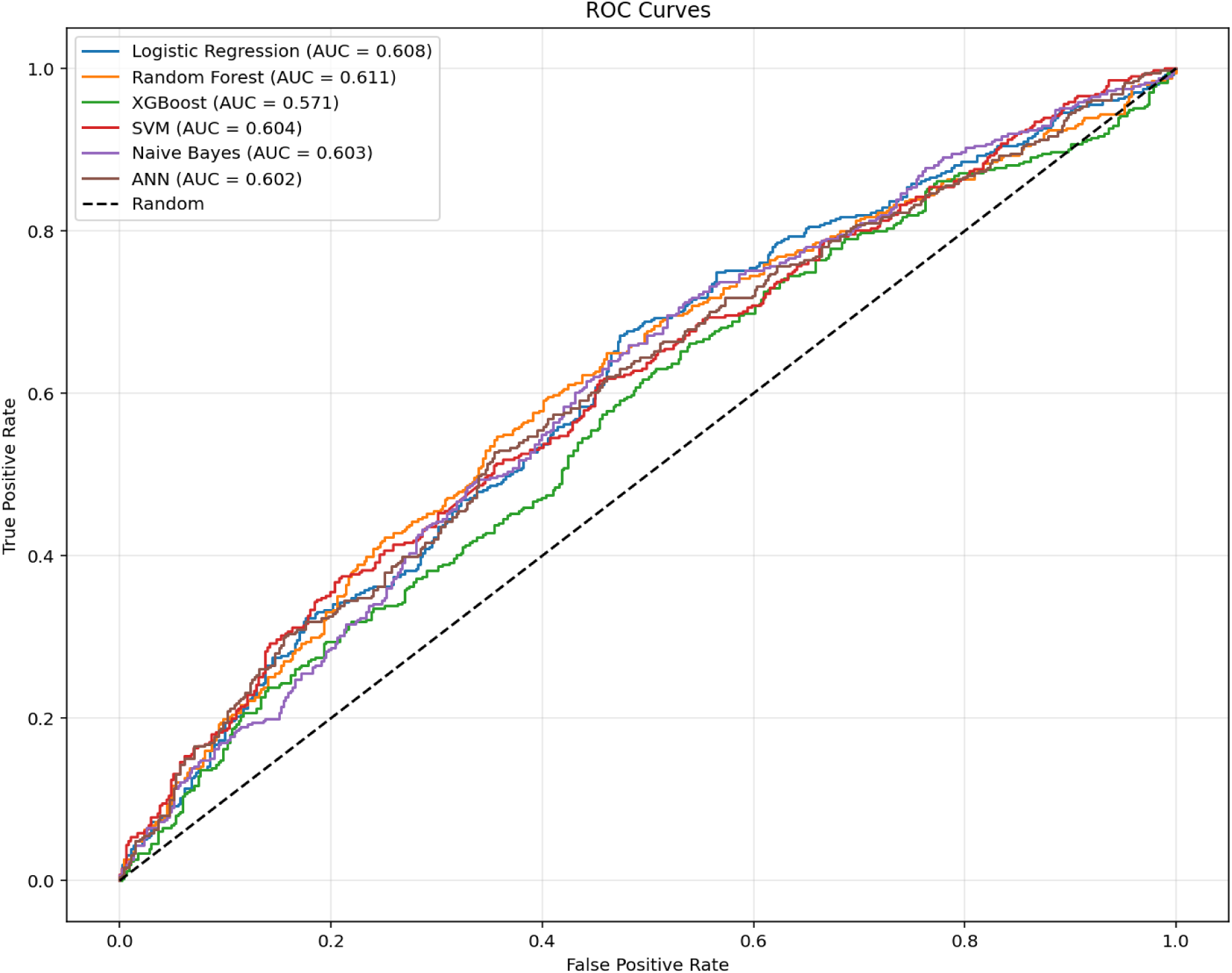
Receiver Operating Characteristic (ROC) Curves for All Models

#### Precision-Recall Curves

The Precision-Recall (PR) curves, shown in Fig 10, provide a complementary evaluation of model performance, particularly relevant given the dataset’s class imbalance (approximately 47% prevalence of tungiasis). Average Precision (AP) scores ranged from 0.529 for XGBoost to 0.576 for SVM. The PR curves demonstrate that precision declined steadily as recall increased for all models. SVM achieved the highest AP (0.576), followed by Random Forest (0.571), Logistic Regression (0.567), ANN (0.566), Naive Bayes (0.557), and XGBoost (0.529). The relatively low AP values indicate limited precision at higher recall levels, suggesting that models struggled to identify tungiasis cases without generating false positives. This pattern is consistent with the class overlap observed in the data and the complex, multifactorial nature of tungiasis transmission.

**Fig 10.**
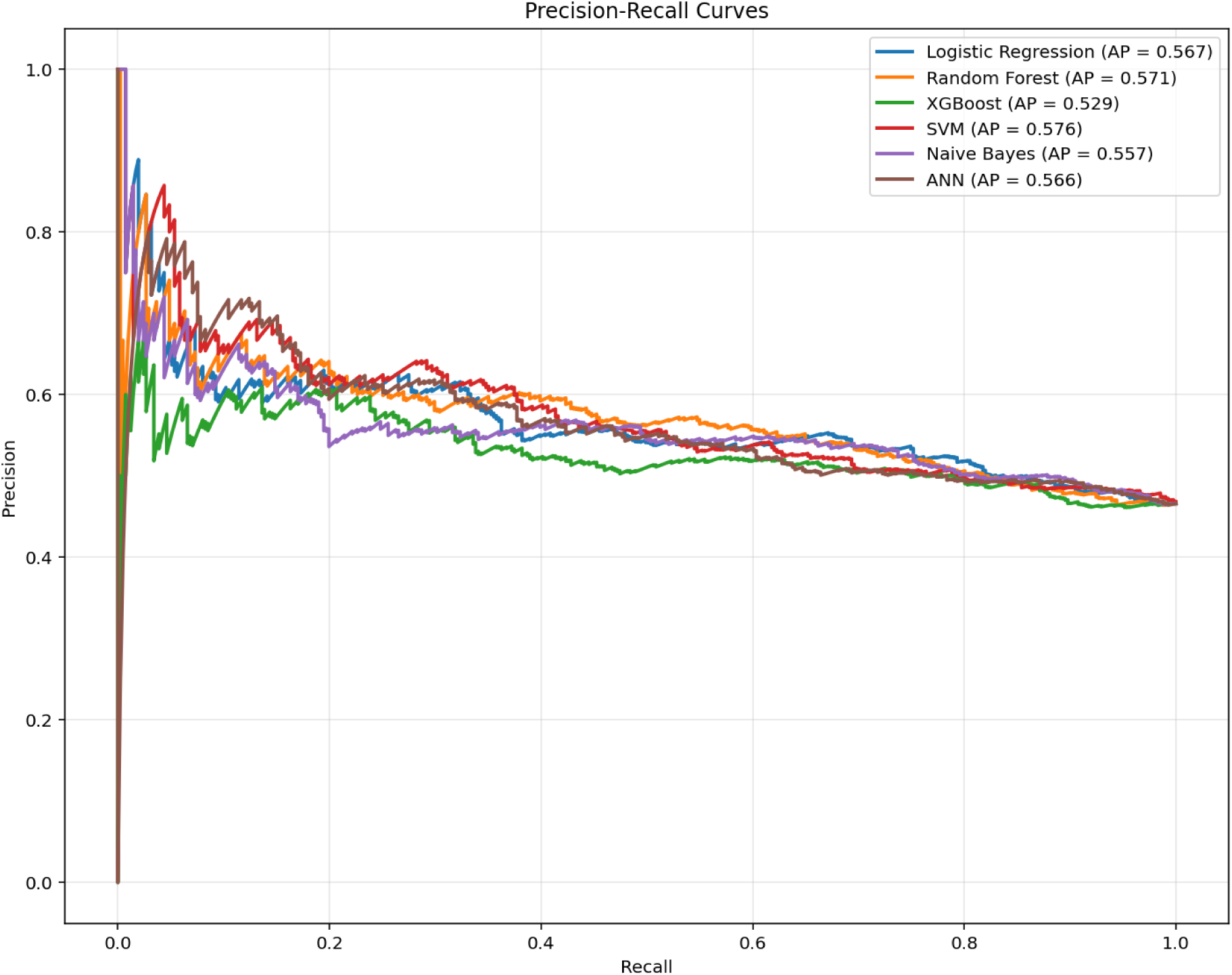
Precision-Recall Curves for All Models

#### Calibration Curves

The calibration curves for all six models are presented in Fig 11. Calibration refers to the agreement between predicted probabilities and observed outcome frequencies; a perfectly calibrated model would produce predicted probabilities that match observed event rates, resulting in points lying on the 45-degree reference line. The results indicate that all models exhibited poor calibration, with predicted probabilities substantially underestimating the true probabilities of tungiasis at most probability ranges. This finding suggests that while the models captured some discriminatory information, the predicted probabilities were not reliable indicators of absolute risk. The poor calibration observed across all models may reflect the inherent complexity of tungiasis transmission and the limited ability of the available predictors to capture the underlying risk factors fully. The calibration results highlight the need for caution when interpreting model outputs as absolute risk estimates, particularly for household-level decision-making.

**Fig 11.**
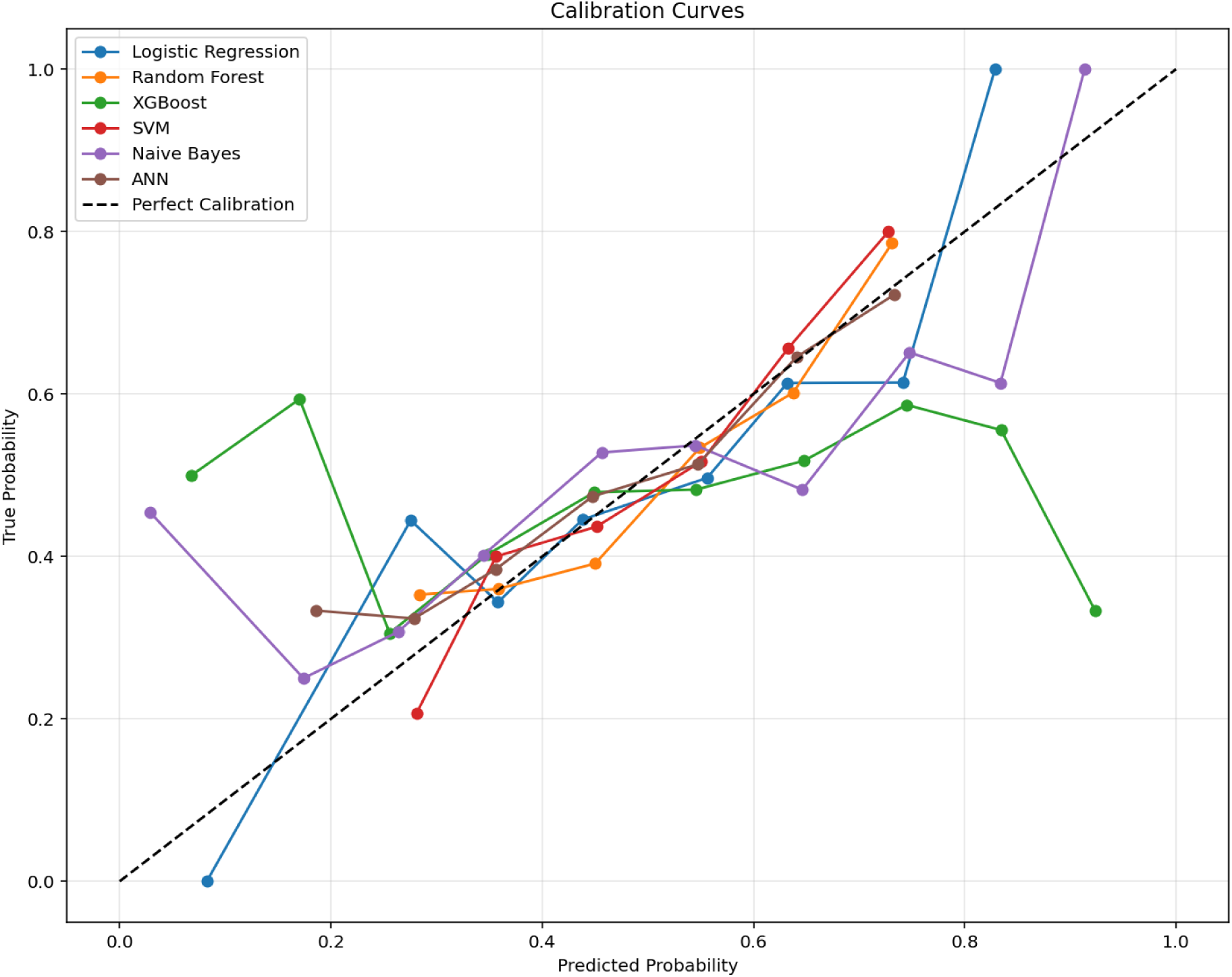
Calibration Curves for All Models

### Scalability Analysis

The scalability analysis provided insights into the computational efficiency of the machine learning models, which is important for potential deployment in resource-constrained settings. Table 7 presents the scalability metrics for all models.

**Table 7.** Scalability Metrics for Machine Learning Models.

| Model | Time (s) | Memory (MB) | Latency (ms) | Size (KB) |
| --- | --- | --- | --- | --- |
| Logistic Regression | 0.032 | 0.516 | 2.298 | 1.233 |
| Random Forest | 2.466 | 0.462 | 15.528 | 2937.900 |
| XGBoost | 0.416 | 0.203 | 3.722 | 225.439 |
| SVM | 0.471 | 0.503 | 20.931 | 169.987 |
| Naive Bayes | 0.012 | 0.538 | 2.779 | 1.596 |

*Note:* Metrics are based on a sample of 1,000 training observations and 100 test observations. Time = training time in seconds; Memory = peak memory usage during training in megabytes; Latency = average inference time per test sample in milliseconds; Size = serialized model size in kilobytes.

**Naive Bayes** demonstrated the highest computational efficiency, with a training time of only 0.012 seconds, negligible memory usage (0.538 MB), and low inference latency (2.779 ms). The model size was also minimal (1.596 KB). These characteristics make Naive Bayes highly suitable for deployment in resource-constrained settings, such as community health facilities with limited computing resources.

**Logistic Regression** also showed excellent efficiency, with training time of 0.032 seconds, low memory usage, and very low inference latency (2.298 ms). The model size was even smaller than Naive Bayes (1.233 KB). The combination of interpretability and computational efficiency makes Logistic Regression an attractive option for rapid deployment and real-time risk assessment.

**XGBoost** demonstrated moderate efficiency, with training time of 0.416 seconds and model size of 225.439 KB. The inference latency was relatively low (3.722 ms), making it suitable for real-time applications. The moderate resource requirements suggest that XGBoost could be deployed on standard computing hardware in health facilities.

**Random Forest** had the highest computational requirements, with training time of 2.466 seconds and model size of 2937.900 KB (approximately 2.9 MB). The inference latency was 15.528 ms, which is higher than other models but still acceptable for most applications. The larger model size reflects the ensemble nature of Random Forest, which stores many decision trees.

**SVM** showed intermediate computational requirements, with training time of 0.471 seconds, model size of 169.987 KB, and the highest inference latency (20.931 ms). The relatively high inference latency may limit SVM’s suitability for applications requiring very fast predictions at scale.

The scalability analysis reveals a clear trade-off between predictive performance and computational efficiency. While Random Forest achieved the highest accuracy (59.4%), it required substantially more computational resources than simpler models. For deployment in resource-constrained settings, Logistic Regression or Naive Bayes may be preferred due to their lower computational requirements, despite their slightly lower predictive performance. For applications where predictive accuracy is prioritized and computational resources are available, Random Forest offers the best performance.

### Feature Importance and Model Interpretability Random Forest Feature Importance

The feature importance analysis based on Random Forest is presented in Fig 12. The results indicate that **Income Log** was the most influential predictor of tungiasis status, with an importance score of 0.057. This was followed by **Rainfall mm** (0.053), **Elevation m** (0.052), **ESI** (Environmental Suitability Index) (0.051), **Soil Moisture** (0.051), and **Humidity pct** (0.051). The top ten features were predominantly environmental and climatic variables, suggesting that environmental conditions play a substantial role in determining the risk of tungiasis. Notably, housing-related variables such as **Floor Type Earthen** (0.032) and behavioral variables such as **Shoe Wearing Rarely** (0.014) also contributed to the model’s predictive performance, although their importance was lower than the environmental variables.

**Fig 12.**
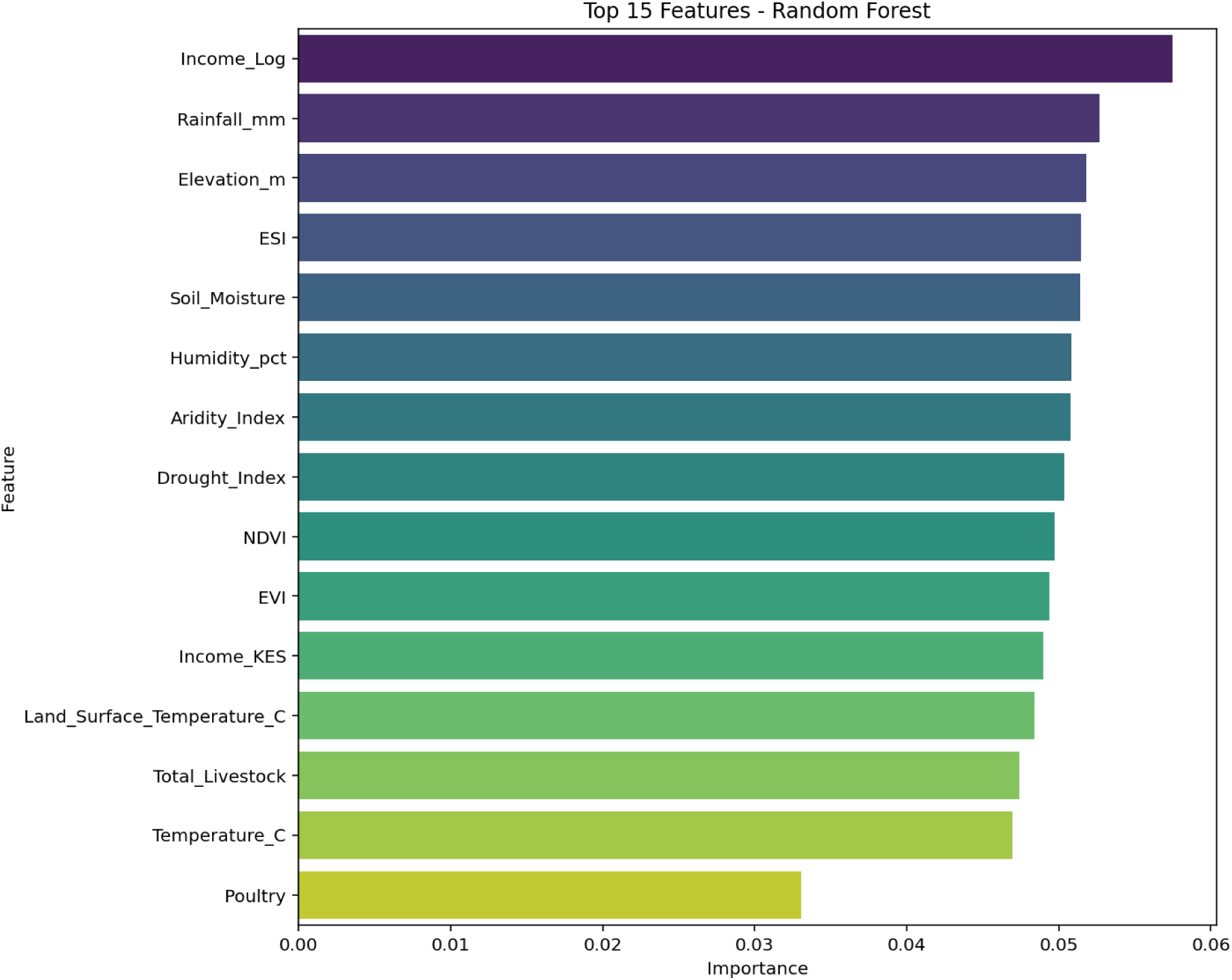
Random Forest Feature Importance

The dominance of environmental and climatic variables in the feature importance ranking is consistent with the ecological nature of tungiasis transmission. *Tunga penetrans* requires specific environmental conditions for survival and reproduction, including adequate soil moisture, temperature, and vegetation cover. The importance of **Income Log** underscores the role of socioeconomic factors in disease risk, as higher-income households may have better housing quality, access to healthcare, and the ability to purchase protective footwear. The relatively low importance of behavioral variables such as shoe-wearing suggests that, while these factors are important, they may be mediated by socioeconomic and environmental conditions.

### Permutation Feature Importance

To validate the Random Forest feature importance results, permutation importance analysis was also conducted (Fig 13). The permutation importance results showed a slightly different ranking, with **Floor Type Earthen** emerging as the most important feature (0.175), followed by **Shoe Wearing Rarely** (0.125). This was followed by **Aridity Index** (0.025), **Income Log** (0.020), **Income KES** (0.018), and **EVI** (0.015). The differences between native Random Forest importance and permutation importance highlight an important distinction: native importance measures how frequently a feature is used in a split. In contrast, permutation importance measures the drop in model performance when a feature’s values are randomly shuffled. The high permutation importance of **Floor Type Earthen** suggests that housing quality is a critical determinant of tungiasis risk, consistent with previous epidemiological studies. The importance of **Shoe Wearing Rarely** underscores the protective effect of footwear, a key behavioral intervention for tungiasis prevention.

**Fig 13.**
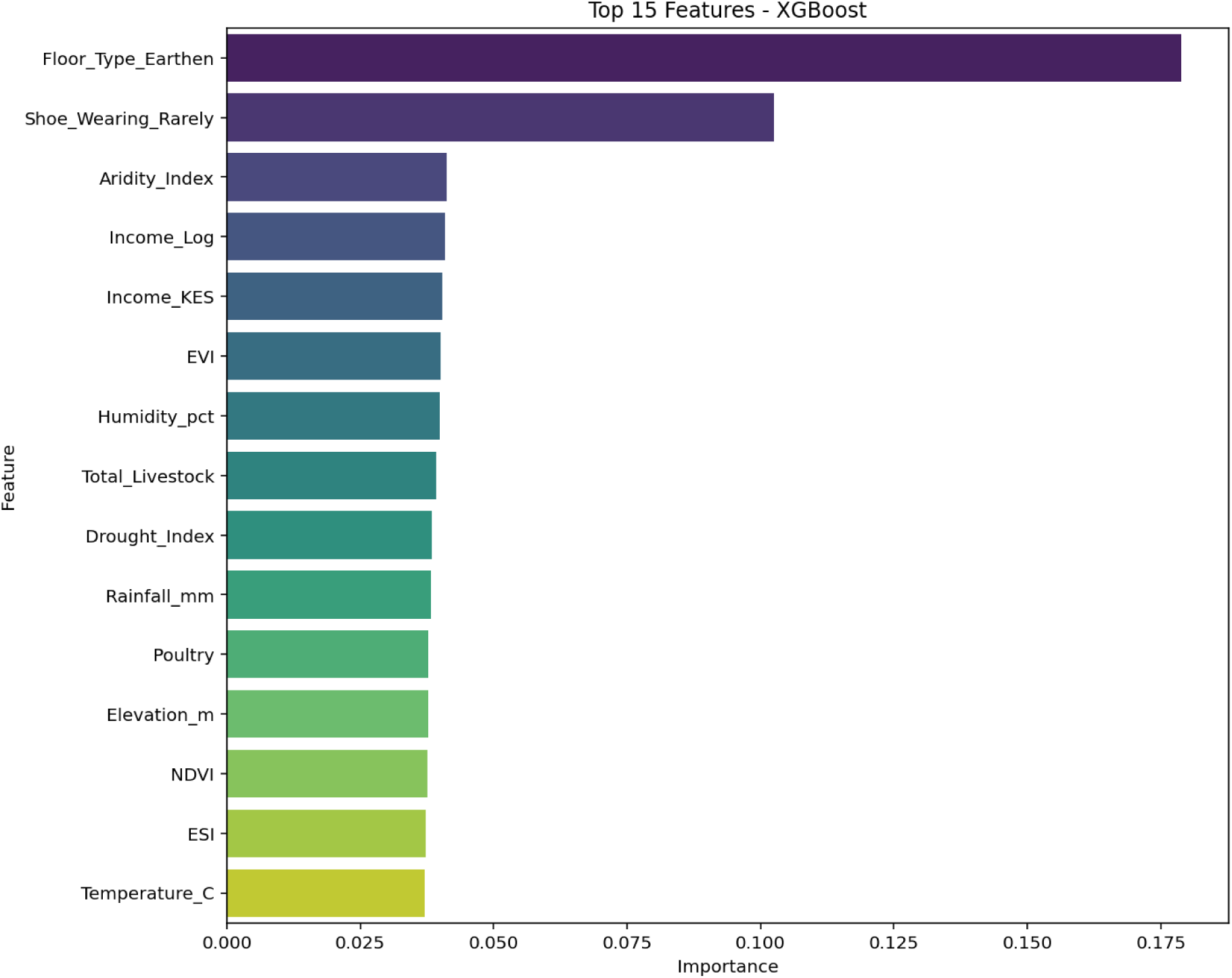
Permutation Feature Importance

### SHAP Analysis

SHAP (SHapley Additive exPlanations) analysis was conducted to provide both global and local interpretations of the Random Forest model predictions. The SHAP interaction plot (Fig 14) illustrates the interaction between **Rainfall mm** and **Income Log**. The plot shows that the effect of **Rainfall mm** on tungiasis risk depends on household income level. For households with low income (blue points), the SHAP values for **Rainfall mm** are more variable, suggesting that rainfall has a stronger effect in low-income households. This interaction pattern highlights the importance of considering both environmental and socioeconomic factors simultaneously when assessing tungiasis risk. The finding suggests that interventions targeting environmental risk factors may be more effective in wealthier households, while low-income households may require additional socioeconomic support.

**Fig 14.**
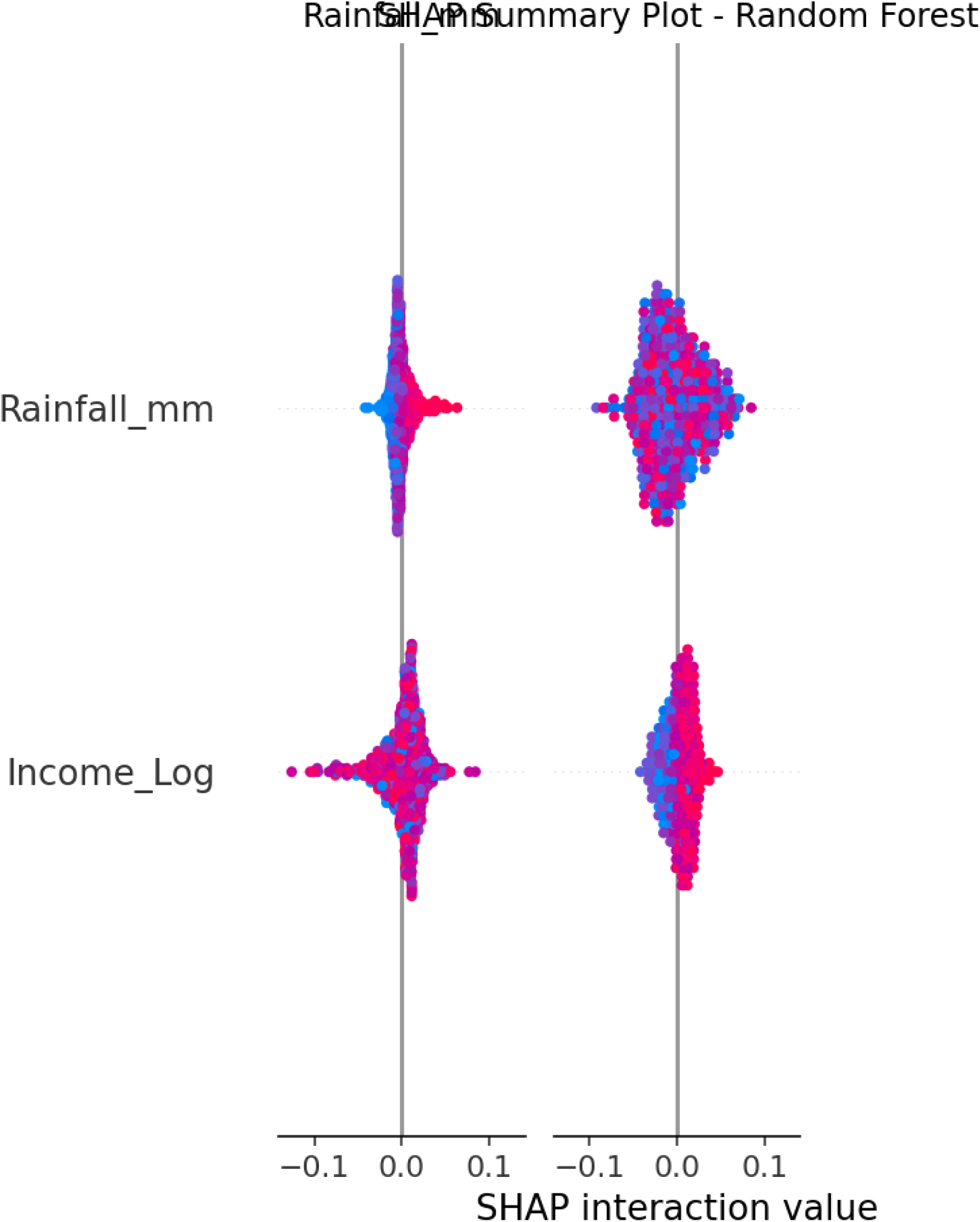
SHAP Interaction Plot: Rainfall mm and Income Log

The SHAP dependence plots for the top four features (Fig 15) reveal the relationship between feature values and SHAP values. For **Income Log**, the plot shows a clear negative relationship: higher income is associated with lower SHAP values (i.e., lower predicted Risk). The relationship appears approximately linear, suggesting that the effect of income on tungiasis risk is consistent across the income distribution. For **Rainfall mm**, the SHAP values show a U-shaped pattern, with both very low and very high rainfall associated with increased Risk. This non-linear relationship may reflect the optimal environmental conditions for *Tungiasis penetrans survival and reproduction at moderate rainfall levels*. For **Elevation m**, higher elevation is associated with lower SHAP values, consistent with the known relationship between altitude and environmental suitability for sand fleas. The **ESI** (Environmental Suitability Index) shows a similar pattern, with higher ESI values associated with lower predicted Risk.

**Fig 15.**
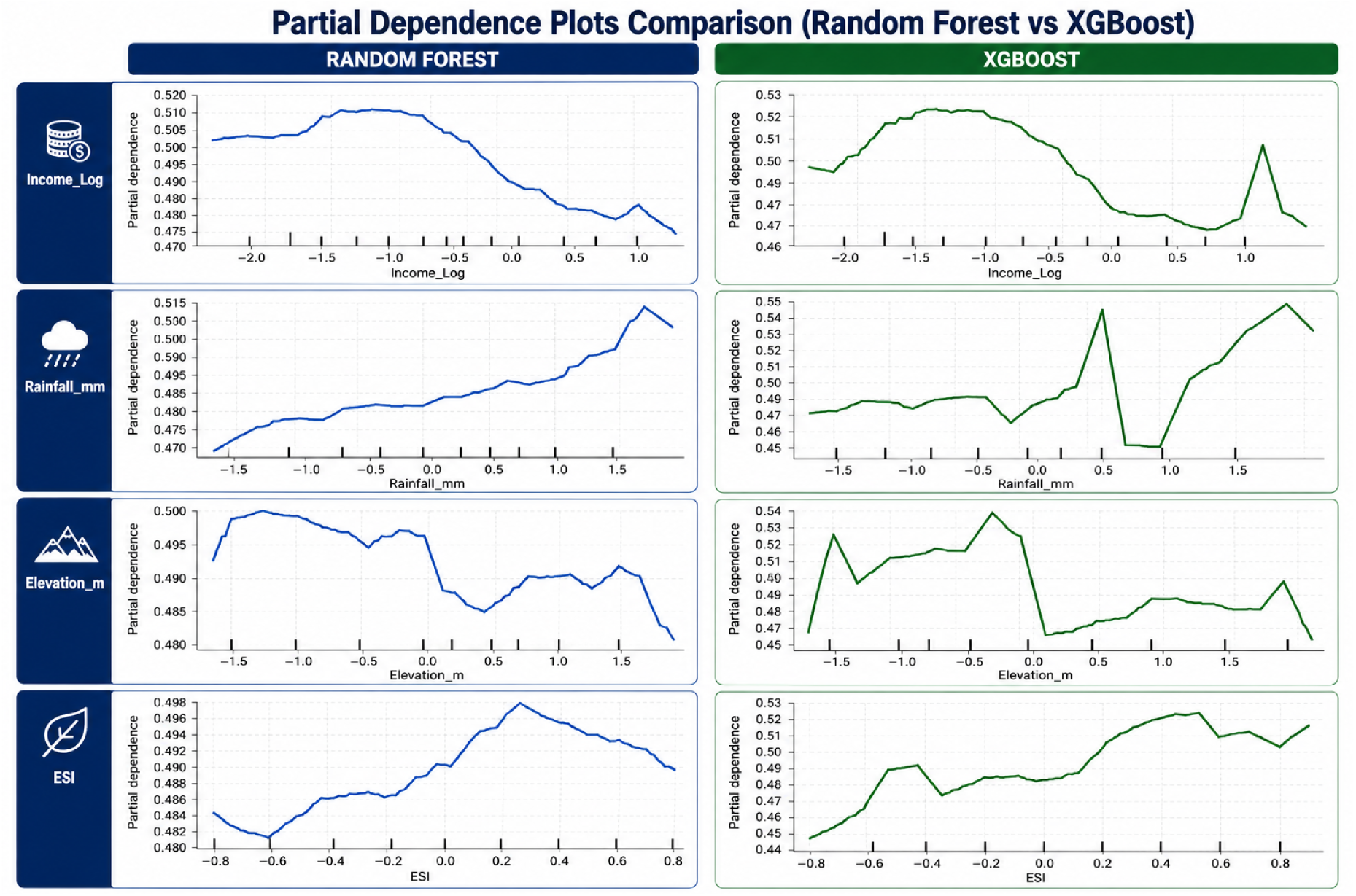
SHAP Dependence Plots for Top Features

#### LIME Explanations

Local Interpretable Model-agnostic Explanations (LIME) were generated for individual predictions to provide insights into the factors driving specific household risk assessments. Three representative instances were selected for explanation, spanning different combinations of actual and predicted outcomes. A summary of the LIME explanations for these instances is presented in Table 8.

**Table 8.**
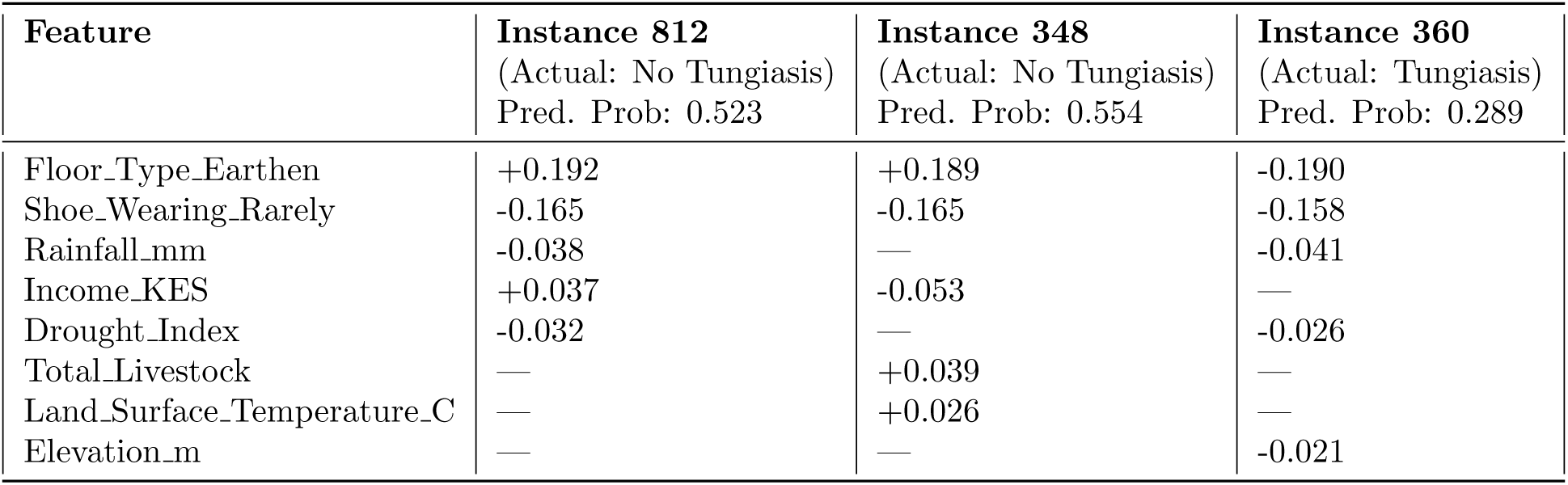
Summary of LIME Explanations for Three Representative Households. The table shows the top contributing features for each instance, with their respective contribution values. Positive contributions increase the predicted probability of tungiasis, while negative contributions decrease it.

| Feature | Instance 812<br>(Actual: No Tungiasis)<br>Pred. Prob: 0.523 | Instance 348<br>(Actual: No Tungiasis)<br>Pred. Prob: 0.554 | Instance 360<br>(Actual: Tungiasis)<br>Pred. Prob: 0.289 |
| --- | --- | --- | --- |
| Floor_Type_Earthen | +0.192 | +0.189 | -0.190 |
| Shoe_Wearing_Rarely | -0.165 | -0.165 | -0.158 |
| Rainfall_mm | -0.038 | — | -0.041 |
| Income_KES | +0.037 | -0.053 | — |
| Drought_Index | -0.032 | — | -0.026 |
| Total_Livestock | — | +0.039 | — |
| Land_Surface_Temperature_C | — | +0.026 | — |
| Elevation_m | — | — | -0.021 |

For the first instance (Fig. 16), the model predicted a 52.3% probability of tungiasis for a household that was actually non-infested. The LIME explanation revealed that the strongest positive contributor was **Floor Type Earthen** (+0.192), indicating that the presence of an earthen floor increased the predicted risk. However, **Shoe Wearing Rarely** (-0.165) strongly decreased the predicted probability, consistent with the protective effect of footwear. **Rainfall mm** also contributed negatively (-0.038), while **Drought Index** had a small negative contribution (-0.032). Interestingly, **Income KES** contributed positively (+0.037), which appears counter-intuitive given that higher income is generally associated with lower disease risk. This unexpected direction may reflect complex interactions among features in this specific household context, or it may indicate that income is acting as a proxy for other unmeasured factors in this particular instance.

**Fig 16.**
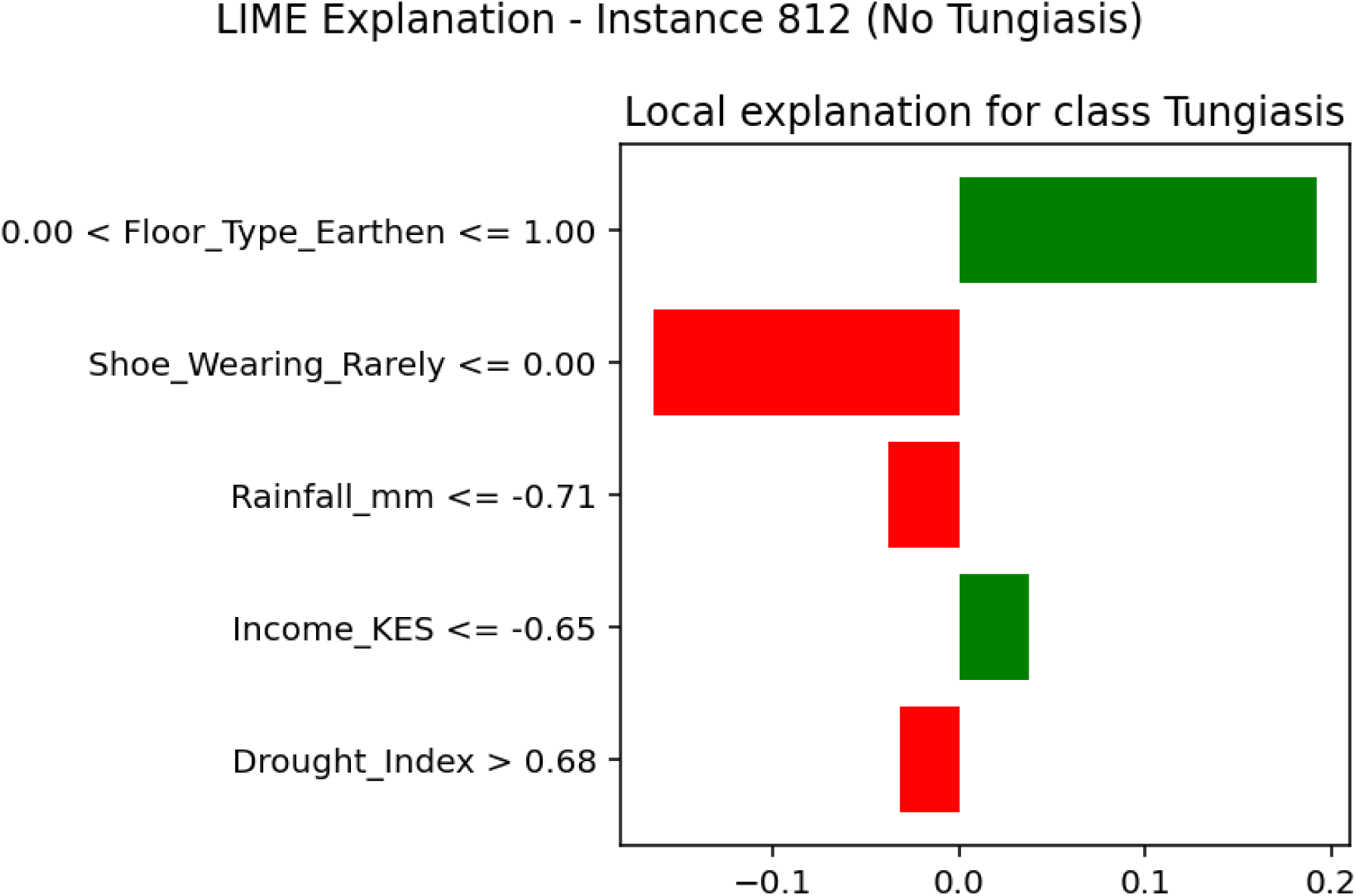
LIME Explanation - Instance 812 (Actual: No Tungiasis).

For the second instance (Fig. 17), the model predicted a 55.4% probability of tungiasis for another non-infested household. The LIME explanation again identified **Floor Type Earthen** (+0.189) as the strongest positive contributor, confirming the epidemiological importance of earthen floors as a risk factor. **Shoe Wearing Rarely** (-0.165) again strongly decreased the predicted probability, reinforcing the protective role of consistent footwear use. However, **Income KES** contributed negatively (-0.053), which aligns with expectations that higher income is associated with lower risk. **Total Livestock** (+0.039) and **Land Surface Temperature C** (+0.026) also contributed positively, suggesting that livestock ownership and warmer land surface temperatures may increase tungiasis risk in certain contexts. This pattern is consistent with established epidemiological evidence: earthen floors provide favorable conditions for *Tunga penetrans*, while livestock may facilitate transmission through close human-animal contact.

**Fig 17.**
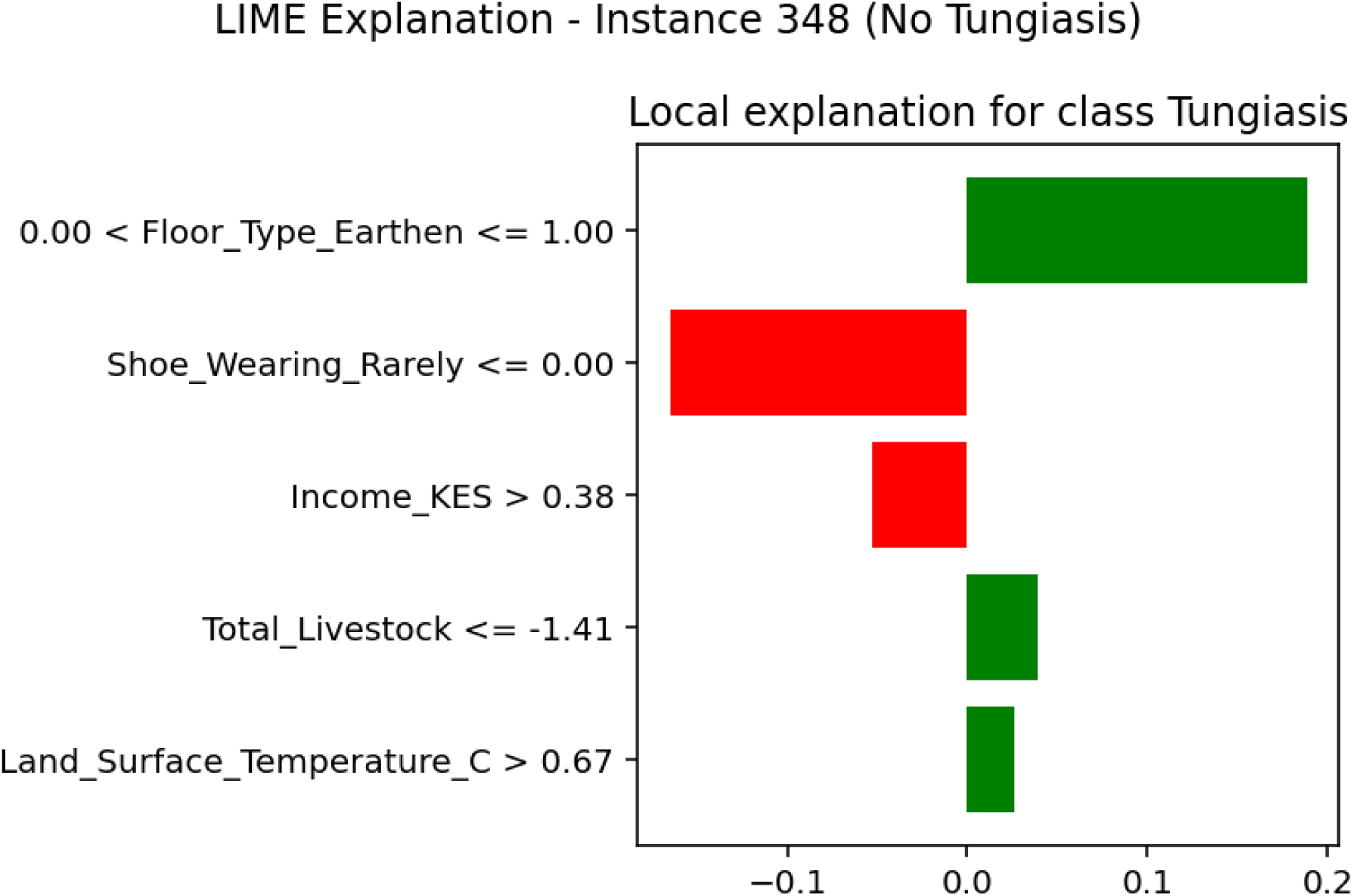
LIME Explanation - Instance 348 (Actual: No Tungiasis).

For the third instance (Fig. 18), the model predicted a 28.9% probability of tungiasis for a household that was actually infested. This represents a case where the model was confidentially incorrect, substantially under-predicting the true risk. The LIME explanation showed that all identified features had negative contributions: **Floor Type Earthen** (-0.190), **Shoe Wearing Rarely** (-0.158), **Rainfall mm** (-0.041), **Drought Index** (-0.026), and **Elevation m** (-0.021). The negative contribution of **Floor Type Earthen** is particularly striking, as the absence of an earthen floor (Floor Type Earthen ≤ 0.00) appears to have reduced the predicted probability, suggesting that the household likely had a cement or other non-earthen floor. However, the presence of **Shoe Wearing Rarely** also contributed negatively, which is counter-intuitive as rare footwear use is generally associated with increased risk. This instance illustrates a clear limitation of the model: important risk factors for this specific household were not captured by the available predictors, leading to a substantial under-prediction of tungiasis risk. Possible unmeasured factors could include household sanitation practices, personal hygiene, treatment-seeking behavior, or other local environmental conditions not included in the dataset.

**Fig 18.**
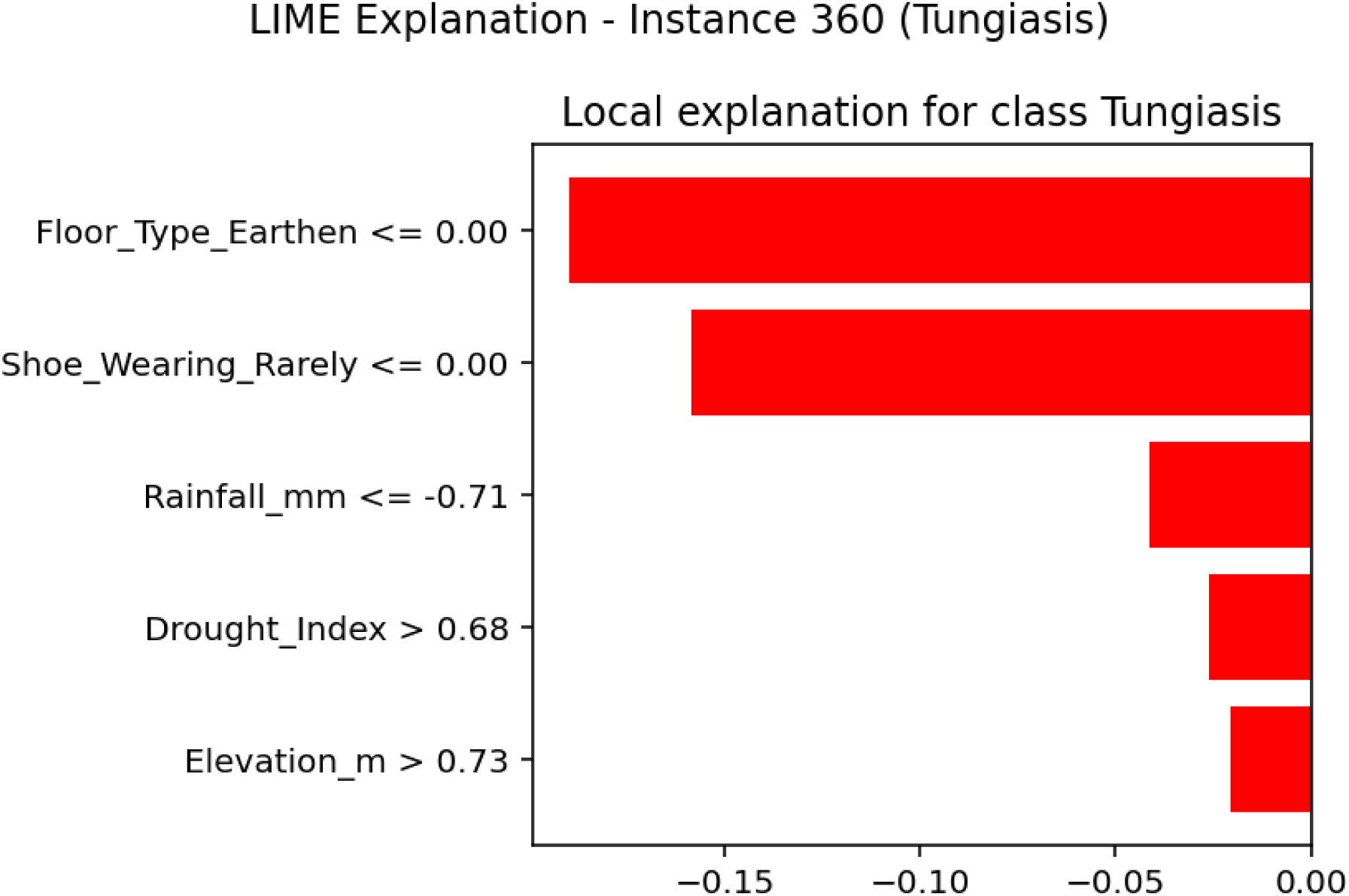
LIME Explanation - Instance 360 (Actual: Tungiasis).

Overall, the LIME explanations highlight the importance of housing quality (particularly earthen floors) and footwear behavior as key determinants of tungiasis risk at the household level. **Floor Type Earthen** consistently emerged as the strongest positive contributor for non-infested households when present, but its absence (or different flooring type) contributed negatively for the infested household that was under-predicted. Similarly, **Shoe Wearing Rarely** consistently decreased predicted probabilities across all instances, reinforcing the protective effect of consistent footwear use—a key behavioral intervention for tungiasis prevention. The variable contributions of environmental and climatic factors (rainfall, drought index, land surface temperature, elevation) across instances reflect the heterogeneous nature of tungiasis risk and the importance of local context.

The case of Instance 360, where the model substantially under-predicted risk for an actually infested household, underscores an important limitation of the current modeling approach. This misclassification suggests that important determinants of tungiasis transmission—such as household sanitation, personal hygiene practices, domestic animal infestation status, occupational exposure, or seasonal variation—were not captured in the available dataset. Improving model performance will require incorporating these additional risk factors in future studies.

### Hotspot Prediction and Risk Mapping

The best-performing model (Random Forest) was used to predict tungiasis risk for all households in the dataset, and the predictions were aggregated at the county level to identify geographic hotspots. Table 9 presents the county-level risk statistics.

**Table 9.**
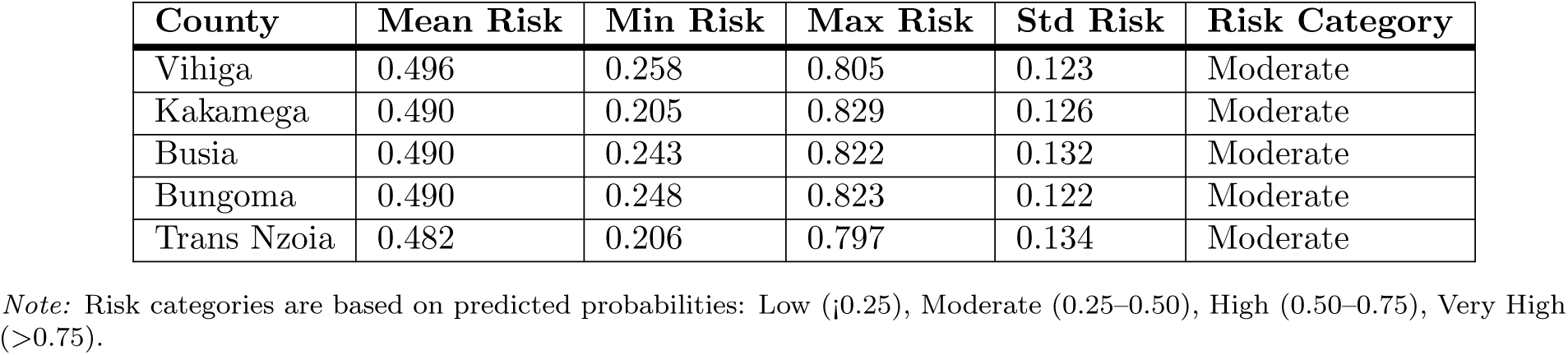
County-Level Tungiasis Risk Predictions.

The county-level risk predictions indicate that all five counties in Western Kenya have moderate average tungiasis risk, with mean probabilities ranging from 0.482 (Trans Nzoia) to 0.496 (Vihiga). While the mean risks were similar across counties, substantial variation was observed within each county, as indicated by the standard deviations ranging from 0.122 to 0.134 and the wide ranges between minimum and maximum household-level risks. This within-county heterogeneity suggests that tungiasis risk is highly localized and may be influenced by household-level factors that vary considerably even within the same geographic area.

Vihiga County had the highest mean Risk (0.496), followed closely by Kakamega, Busia, and Bungoma (all around 0.490). Trans Nzoia had the lowest mean Risk (0.482). However, the relatively small differences in mean Risk across counties suggest that all five counties face substantial tungiasis burden and require targeted interventions. The maximum household-level Risk exceeded 0.80 across all counties, indicating that some households have very high predicted tungiasis risk despite the county-level averages being moderate.

The predicted risk maps (Fig 19) provide a visual representation of the spatial distribution of tungiasis risk across the study area. The maps reveal localized risk patterns, with some sub-locations showing consistently higher predicted Risk than others. These high-risk areas likely correspond to clusters of households with high levels of key risk factors, such as earthen floors, low income, infrequent shoe-wearing, and unfavorable environmental conditions. The identification of these localized risk clusters can guide the targeting of interventions, such as health education campaigns, provision of footwear, and housing improvement programs.

**Fig 19.**
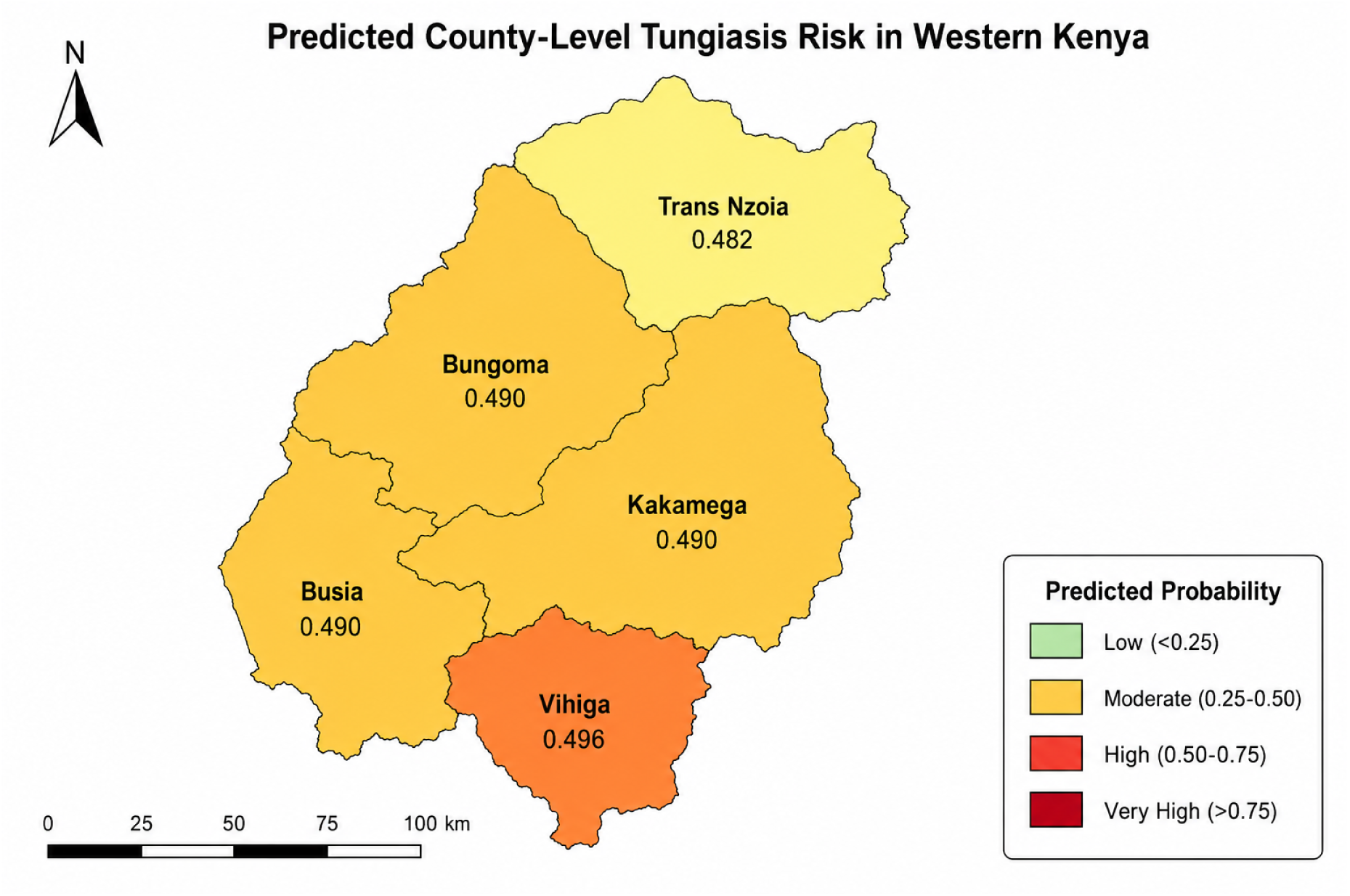
Predicted Tungiasis Risk Map by County

The finding that Risk is localized rather than uniformly distributed across counties has important implications for public health planning. While county-level averages may appear similar, the underlying spatial heterogeneity suggests that interventions should be targeted at high-risk sub-locations rather than applied uniformly across entire counties. This targeted approach would be more cost-effective and likely more effective in reducing the burden. The risk maps developed in this study provide a practical tool for decision-makers to prioritize areas for intervention and allocate resources more efficiently.

## Discussion

This study evaluated the use of machine learning integrated with explainable artificial intelligence (XAI) to predict household-level tungiasis risk in Western Kenya. Overall, the findings demonstrate that interpretable machine learning provides a useful framework for identifying communities at elevated risk while revealing the environmental, socioeconomic, housing, and behavioral factors that drive disease transmission. Although all evaluated models achieved only moderate predictive performance, they consistently identified meaningful risk patterns, indicating that machine learning can complement conventional epidemiological approaches in guiding surveillance and targeted interventions.

The observed tungiasis prevalence of 46.6% confirms that the disease remains a substantial public health problem in Western Kenya, consistent with previous reports from endemic regions [9, 11]. Marked differences in prevalence between counties further demonstrate that transmission is spatially heterogeneous rather than uniformly distributed. Such variation likely reflects differences in environmental conditions, housing quality, socioeconomic status, sanitation, and access to healthcare [9, 10, 14]. These findings reinforce the importance of geographically targeted interventions instead of uniform county-wide control strategies.

The identified risk factors highlight the close relationship between tungiasis and poverty. Households with earthen floors, lower income, lower educational attainment, farming occupations, and inconsistent footwear use experienced a greater disease burden than households with improved housing and socioeconomic conditions. These findings are consistent with previous epidemiological studies demonstrating that poor housing provides favorable conditions for the development of *Tunga penetrans*, while regular footwear use reduces exposure to infested soil [1, 10, 13]. Together, these results emphasize that sustainable tungiasis control requires improvements in housing, socioeconomic conditions, and health education alongside routine treatment programs.

A key contribution of this study is the integration of machine learning with explainable artificial intelligence. While previous studies have primarily relied on conventional statistical methods to identify tungiasis risk factors [9–11, 14], machine learning provides greater flexibility for modelling nonlinear relationships and complex interactions among predictors. The incorporation of SHAP and LIME further improves model transparency by identifying the variables driving individual and overall predictions, thereby increasing confidence in the use of these models for public health decision-making.

Despite these methodological advantages, predictive performance remained moderate across all evaluated algorithms, with Random Forest providing only modest improvements over simpler approaches. This finding suggests that important determinants of tungiasis transmission were not fully represented in the available dataset. Variables such as household sanitation, domestic animal infestation, seasonal variation, occupational exposure, and detailed behavioral practices may explain additional variability and improve future predictive performance. Consequently, the principal value of the proposed framework lies not only in prediction accuracy but also in its ability to identify high-risk communities and improve understanding of disease drivers.

The explainability analyses were consistent with established epidemiological knowledge. SHAP identified household income, rainfall, elevation, environmental suitability, and soil moisture as the most influential predictors, while LIME consistently highlighted housing quality and footwear behavior in individual predictions. These complementary findings strengthen confidence in the developed models and demonstrate that explainable AI can bridge the gap between predictive analytics and actionable public health evidence.

Environmental factors also played an important role in determining tungiasis risk. Rainfall, elevation, humidity, soil moisture, and environmental suitability consistently influenced model predictions, reflecting the ecological requirements of *Tunga penetrans* for survival and reproduction [13, 32]. The observed nonlinear relationships, particularly for rainfall, illustrate the ability of machine learning to capture complex environmental interactions that are often difficult to represent using conventional regression models.

### Spatial heterogeneity and implementation

The generated risk maps revealed substantial variation within counties, indicating that tungiasis is concentrated in localized hotspots rather than being evenly distributed. This spatial clustering is consistent with previous reports from endemic settings [13, 14]. Consequently, predictive risk mapping offers a practical framework for prioritizing surveillance, community education, environmental improvement, and treatment in areas where they are most needed. Integrating SHAP and LIME further enhances implementation by providing transparent explanations that can increase confidence among public health practitioners and support evidence-based decision-making.

Although Random Forest achieved the highest predictive performance, Logistic Regression and Naive Bayes produced comparable results while requiring substantially fewer computational resources. These simpler algorithms may therefore provide practical alternatives for deployment in resource-constrained settings. Nevertheless, all models exhibited poor calibration, indicating that predicted probabilities should be interpreted primarily as relative measures of risk rather than precise estimates of disease probability. Future work should prioritize probability calibration before operational deployment.

### Strengths and limitations

A major strength of this study is the integration of multiple machine learning algorithms with explainable artificial intelligence using demographic, socioeconomic, behavioral, housing, and environmental data.

Rigorous model development procedures, including stratified sampling, cross-validation, multiple performance metrics, and explainability analyses, improve the robustness and reproducibility of the findings. The generated risk maps further demonstrate the practical potential of predictive modelling to support disease surveillance and intervention planning.

Several limitations should be acknowledged. First, the study relied on secondary cross-sectional data, limiting causal inference and introducing the possibility of measurement error. Second, the analysis was restricted to Western Kenya, which may limit generalizability to other endemic regions. Third, the moderate predictive performance suggests that important determinants of tungiasis, including household sanitation, domestic animal infestation, seasonal dynamics, and other behavioral factors, were unavailable in the dataset. Finally, although SHAP and LIME improved model interpretability, the proposed framework has not yet undergone external validation or prospective evaluation and should therefore be regarded as a decision-support framework rather than a fully operational surveillance system.

### Implications for policy and future research

The findings demonstrate that interpretable machine learning has considerable potential to strengthen tungiasis surveillance by identifying communities at elevated risk and informing targeted interventions. Housing improvement, promotion of consistent footwear use, environmental management, and community health education should remain central components of integrated control strategies. Because disease risk is influenced by interacting environmental and socioeconomic factors, effective control will require coordinated action across the health, housing, education, agriculture, and environmental sectors.

Future research should incorporate additional environmental, behavioral, and household variables, improve probability calibration, and validate the proposed framework using independent datasets from multiple endemic settings. Longitudinal studies would further support prediction of seasonal transmission patterns and facilitate the development of early warning systems for tungiasis surveillance and control.

## Conclusion

This study evaluated the application of machine learning integrated with explainable artificial intelligence to predict household-level tungiasis risk in Western Kenya. Among the evaluated algorithms, Random Forest achieved the highest overall predictive performance, while SHAP and LIME enhanced model interpretability by identifying the environmental, socioeconomic, housing, and behavioral factors that contributed most to model predictions.

The findings indicate that tungiasis risk is influenced by complex interactions among environmental and socioeconomic determinants and demonstrate that interpretable machine learning can complement conventional epidemiological approaches for identifying populations and locations at relatively higher risk. Although predictive performance was moderate, the explainability analyses provide useful insights into the key drivers of disease risk and highlight the potential value of machine learning for supporting evidence-informed surveillance and intervention planning.

Future research should incorporate additional behavioral, environmental, and household-level predictors, improve model calibration, and validate the proposed framework using independent datasets from diverse endemic settings. Collectively, these findings provide a reproducible foundation for further development and evaluation of interpretable machine learning approaches for tungiasis surveillance and public health decision support.

## Data Availability

The data supporting the findings of this study have been deposited in Zenodo and are publicly available at: https://doi.org/10.5281/zenodo.21391066

## Acknowledgments

The authors sincerely thank the Kenya Medical Research Institute (KEMRI), Kisumu Centre, for granting access to the secondary dataset used in this study. We also acknowledge the researchers, field staff, and study participants whose efforts in data collection made this research possible. Their contribution has been invaluable in advancing research on tungiasis and supporting the development of data-driven approaches for disease risk prediction.

